# Influence of Poor Sleep on Cardiovascular Disease-Free Life Expectancy: A Multi-Resource-Based Population Cohort Study

**DOI:** 10.1101/2022.10.27.22281630

**Authors:** Bo-Huei Huang, Borja del Pozo Cruz, Armando Teixeira-Pinto, Peter A. Cistulli, Emmanuel Stamatakis

## Abstract

**Background:** The complexity of sleep hinders the formulation of sleep guidelines. Recent studies suggest that different unhealthy sleep characteristics jointly increase the risks for cardiovascular disease (CVD). This study aimed to estimate the differences in CVD-free life expectancy between people with different sleep profiles.

**Methods:** We included 308683 middle-aged adults from the UK Biobank among whom 140181 had primary care data linkage. We used an established composite sleep score comprising self-reported chronotype, duration, insomnia complaints, snoring, and daytime sleepiness to derive three sleep categories: poor, intermediate, and healthy. We also identified three clinical sleep disorders captured by primary care and inpatient records within two years before enrollment in the cohort: insomnia, sleep-related breathing disorders, and other sleep disorders. We estimated sex-specific CVD-free life expectancy with three-state Markov models conditioning on survival at age 40 across different sleep profiles and clinical disorders.

**Results:** We observed a gradual loss in CVD-free life expectancy toward poor sleep such as, compared with healthy sleepers, poor sleepers lost 1·80 [95% CI 0·96-2·75] and 2·31 [1·46-3·29] CVD-free years in females and males, respectively, while intermediate sleepers lost 0·48 [0·41-0·55] and 0·55 [0·49-0·61] years. Among men, those with clinical insomnia or sleep-related breathing disorders lost CVD-free life by 3·84 [0·61-8·59] or 6·73 [5·31-8·48] years, respectively. Among women, sleep-related breathing disorders or other sleep disorders were associated with 7·32 [5·33-10·34] or 1·43 [0·20-3·29] years lost, respectively.

**Conclusions:** Both self-reported and doctor-diagnosed poor sleep are negatively associated with CVD-free life, especially pronounced in participants with sleep-related breathing disorders.

## Background

The World Health Organization has recognized sleep as a critical health state and health-related behavior.(1) Although one-fourth of Europeans have insomnia,(2) with very few exceptions,(3) there is very little national guidance on sleep, reflecting the complexity of sleep health as a multi-dimensional behavior(4) as well as the lack of longitudinal evidence base on population-wide effects of poor sleep.(3,5)

One major difficulty in evaluating the health impacts of sleep is its complexity as a biological process and behavior.(4) Current estimates of optimal nocturnal sleep range in adults is seven to less than nine hours per night,(6,7) with little evidence-based guidance on other health-related sleep characteristics, such as quality and timing.(5,8) While most cohorts fail to capture the multi-dimensional nature of sleep, recent studies have highlighted joint effects of different sleep characteristics with risks for cardiovascular disease (CVD).(8,9) With a composite sleep score comprising chronotype, duration, insomnia complaint, snoring, and daytime sleepiness, Fan et al. found that healthy sleepers had a 35% lower risk of incident CVD than poor sleepers.(9) Using the same set of sleep measures, our previous work further suggested that poor sleepers had a 39% higher risk of CVD mortality than healthy sleepers.(8) These findings indicated that a traditional mutually adjusted approach could not capture the multiplicative effects of different sleep characteristics, and the derived individual/independent effect of a single characteristic could be spurious.

Compared to ratio-based estimation, years of life lost free of diseases, the difference in disease-free life expectancy between people with different characteristics provides a tangible metric on the burdens of exposures taking both morbidity and mortality into account. Life expectancy is easy to communicate to policymakers, media, and the general population.(10–12) However, no study has used such measures to evaluate the health burden of comprehensive sleep characteristics. Primary care (general practice) records have distinct advantages in capturing incident sleep disorders more comprehensively and objectively.(5)

To our knowledge, no prior sleep research has used primary care diagnoses to estimate the health burden of the compromised life expectancy of sleep disorders. Therefore, this study aimed to estimate CVD-free years of life lost among people with poor sleep, evaluated by self-report and clinical primary care diagnosis.

## Methods

### Data Source

We retrieved individual data from the UK Biobank, a population-based prospective cohort that recruited participants in 22 assessment centers throughout England, Wales, and Scotland between 2006 and 2010 (https://www.ukbiobank.ac.uk/). Full cohort details can be found elsewhere.(13,14) In brief, more than half a million participants (response rate 6%) aged 40 to 69 provided informed consent to enrolment and data linkage to electronic health records. The baseline assessment included self-reported questionnaires, a computer-assisted personal interview, physical examinations, and biological sampling. The UK Biobank keeps removing all records of dropping out participants, so retrieving loss of follow-up is inapplicable. By the time we conducted the current study (October 2021), there were 502,459 participants with data available. The National Health Service and the National Research Ethics Service have approved the UK Biobank (Ref. 11/NW/0382).

### Electronic Health Records

The UK Biobank repository accesses national registered data on hospital inpatient diagnoses and death for all participants across the UK through the National Health Service (NHS). The NHS encoded records with the International Classification of Diseases (ICD), of which both the Ninth Revision (ICD-9) and the Tenth Revision (ICD-10) were available for the inpatient records, while only ICD-10 was applied to the death registry. Inpatient records were availed from 1981 onwards. Both registries were censored up to March 2021, with slight differences between countries (**appendix pp 2**).

Primary care data on clinical events and prescriptions covered 227834 participants. Primary care clinical events were coded with Read codes version 2 (Read v2) or Clinical Terms Version 3 (CTV3), while prescriptions were mainly coded with Read v2, British National Formulary (BNF), or Dictionary of Medicines and Devices (dm+d). Further details of primary care data sources, cleaning, and harmonization are provided in **appendix pp 2, 19**. The primary care was censored to September 2018, with data availability varied between participants and was determined by the registration date with a primary care practice.

### CVD Incidence and All-Cause Mortality

We identified the incidence of hospital inpatient disease events based on ICD-9 and ICD-10: CVD (I01 to I09, I11, I13, I21 to I25, I27, I3, I4, I50, I51, I6, I7, I80 to I84, I87 to I9, R54).(10,15) The corresponding ICD-9 was provided in **appendix pp 3**. The date of events and death were retrieved from the NHS electronic health records described above.

### Sleep Exposure

As part of the baseline questionnaire, participants reported five sleep characteristics, based on which we further identified five healthy phenotypes, including no usual insomnia complaints, adequate sleep duration (7 to <9 hr/d), no snoring, morning chronotype, and no frequent daytime sleepiness.(9) We scored participants from 0 to 5, according to the count of healthy characteristics and categorized them into three groups: “healthy sleep” (≥ 4 composite sleep score); “intermediate sleep” (2 or 3 score); and “poor sleep” (≤ 1 score). This categorization has been proven to distinguish different CVD risk profiles, and the simple addition approach showed a similar predictive power to a weighted score.(9) The original questions and options were provided in **appendix pp 3**. These definitions and scores have shown excellent convergent validity with CVD incidence and mortality.(8,9)

We identified recent clinical sleep disorder events (two years before enrolment) based on inpatient admissions, primary care clinical events, and sleep disorder-specific prescriptions (BNF Chapter 4 Section 1: hypnotics and anxiolytics). Modified from the definition provided by the American Sleep Association and American Academy of Sleep Medicine,(16,17) we distinguished five different sleep disorders, including insomnia, hypersomnia, sleep-related breathing disorders, circadian rhythm sleep disorders, and parasomnias (including sleep-related bruxism). In addition, we grouped hypersomnia, circadian rhythm sleep disorders, parasomnias, non-specific sleep disorders (*e*.*g*., “poor sleep pattern”), and sleep medication prescriptions without a corresponding clinical event into “other sleep disorders.” We provided detailed codes for each classification system in **appendix pp4-7**. Since the clinical diagnosis of sleep disorders mainly refers to more than one self-reported sleep characteristic, we did not further integrate both self-reported and clinically unhealthy sleep.

## Statistical analysis

We set the follow-up from the UK Biobank enrollment to the date of death, inpatient or death registry censoring, or the 81^st^ birthday, whichever came first. To avoid overestimation, we constrained the censoring age (τ restriction) at 81, corresponding to the oldest age of death in the analyzed sample.(18) Considering the pathophysiological differences in CVD risks between sexes,(19) we stratified all our models by sex. To evaluate the sex-specific CVD-free residual lifetime of participants with different exposures, we applied continuous-time three-state Cox Markov survival models.(20,21) All participants started with the CVD-free state and subsequently either (a) maintained healthy until censored, (b) were diagnosed with the disease(s) and lived until censored, (c) maintained healthy then deceased, or (d) were diagnosed with the disease(s) then deceased. We estimated state-specific residual lifetime at age 40,(22) and calculated the total and CVD-free years of life lost by comparing the marginal life expectancies spent in a healthy state between the population with questionnaire-based poor/intermediate sleep (exposure) and those with healthy sleep (reference), or between the population with each diagnosed sleep disorder (exposure) and those without (reference). Based on *a priori* defined directed acyclic graphs (**appendix pp 20**),(23) we selected the following potential covariates: age, socioeconomic status, mental health issues, perceived health, body mass index (BMI), economic activity, and shift work, cigarette smoking, alcohol consumption, diet quality, discretionary screen time, and physical activity. The detailed definitions and the original questions of covariates were provided in **appendix pp 8**. Models examining each self-reported sleep characteristic or diagnosed sleep disorder were further mutually adjusted for the remaining characteristics or disorders. For all the life expectancy estimations, we applied median/ mean value for categorical/numerous covariates, and for estimations of each self-reported sleep characteristic/clinical sleep disorder, the remaining characteristics or disorders were set to healthy.

We obtained the 95% CIs for life expectancy estimations via nonparametric bootstrapping with 1000 iterations. All data were processed in SAS 9·4 (on secured local computers), and analyses were performed on R 4·1·1 (on secured local computers) or R 4·0·4 (on the University of Sydney’s high performance computing cluster Artemis). To improve comparability, we repeated all analyses with Fine-Gray sub-distribution hazards models, in which non-CVD death was entered as a competing risk to incident CVD.(24) Covariates stayed the same as in the primary analyses.

## Results

We sequentially excluded participants with missing data in any of the five self-reported sleep characteristics (n = 91802), with an inpatient or self-reported history of CVD (n = 54703), with missing covariates (n = 32765), and with incident CVD or fatal events within the first two years of follow-up (n = 14506), leaving 308683 participants for the core analysis of self-reported and 140181 participants for the clinically confirmed sleep events data (**Figure 1, appendix pp 9-11**). Among the 308683 participants, with a mean follow-up of 11·8 (1·3) years from a mean age 56·2 (8·1), there were 13790 deaths from any cause and 53064 incident CVD, corresponding to 3653450 person-years at risk of death (**Table 1**). At baseline, 60% of the participants reported healthy sleep, followed by 38% and 2% reporting intermediate and poor sleep, respectively (**Table 1, appendix pp 12-13**). Participants who suffered from mental health problems, perceived poor health, consumed a low-quality diet, had high BMI, faced higher socioeconomic deprivation, retired, smoked/drank more, spent more discretionary time on screen, and exercised less tended to have poor sleep in both sexes. Women were more likely to report healthy sleep despite no appreciable differences in corresponding diagnoses (**appendix pp 14-15**).

**Table 1.**
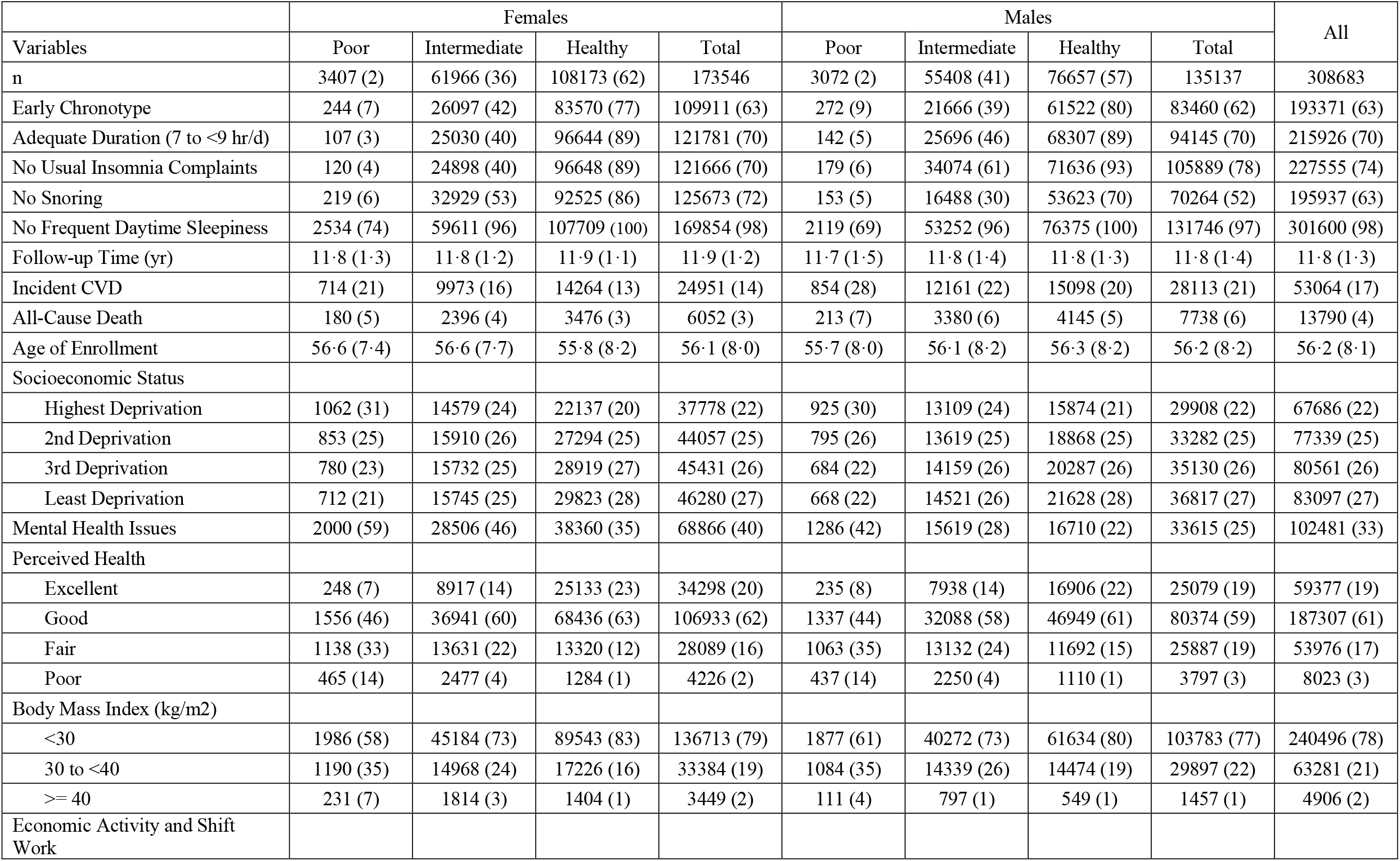
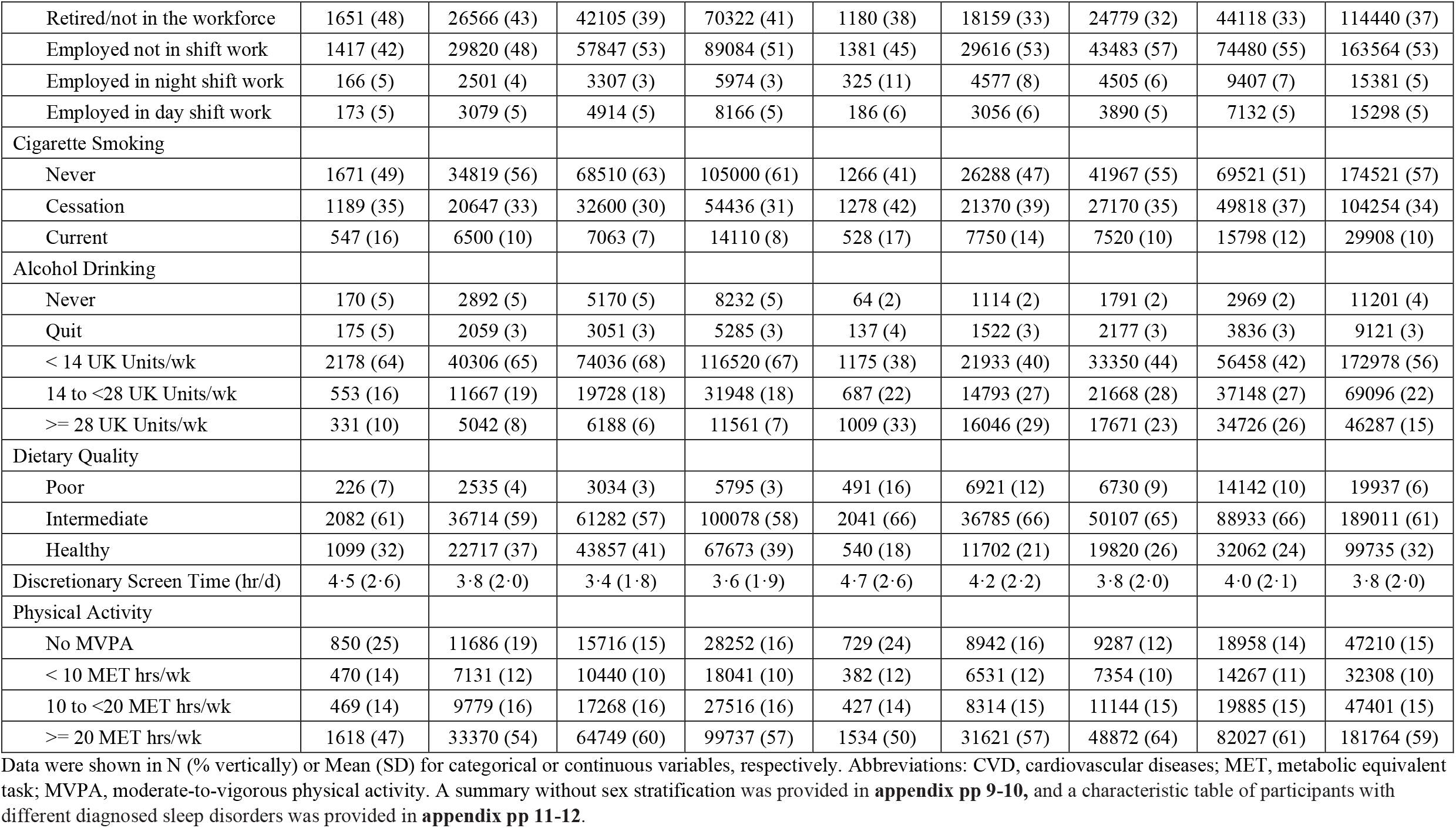
The basic characteristics of participants with different questionnaire-based sleep characteristics

**Figure 1.**
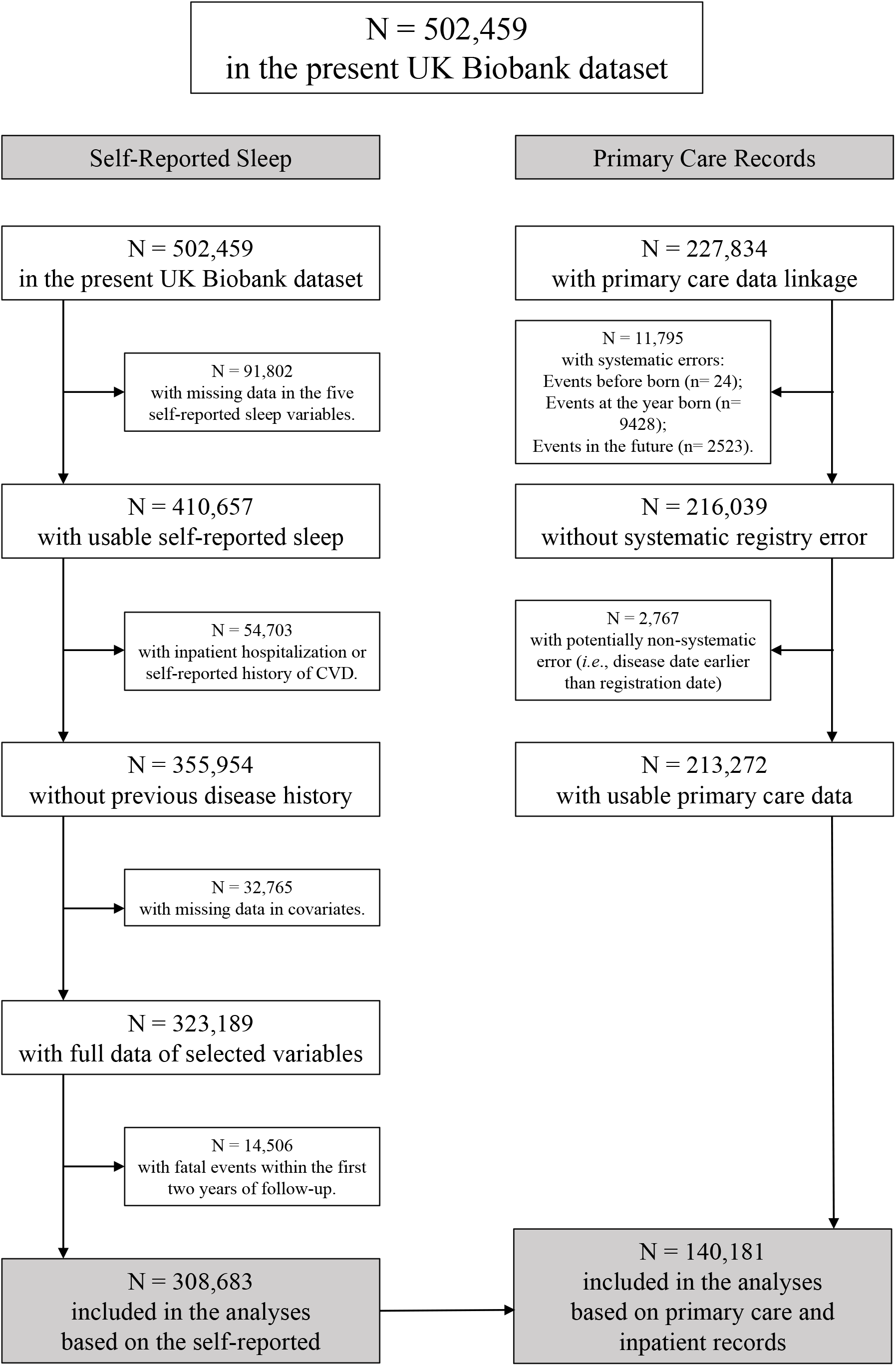
The participants’ exclusion flowchart Abbreviations: CVD, cardiovascular diseases. The detailed missing pattern was provided in **appendix pp 16-18**.

At age 40, total life expectancy was an additional 39·07 [38·95-39·17] and 38·29 [38·16-38·41] years (**appendix pp 16**), while CVD-free life expectancy was 33·05 [32·91-33·21] and 30·03 [29·88-30·20] years for females and males (**Table 2**). Although total life expectancy did not differ between sleep groups (**Figure 2A, appendix pp 17**), there was a gradual decrease in CVD-free life expectancy toward a higher number of self-reported sleep characteristics in both sexes (**Table 2, Figure 2A**). Women with poor sleep had a CVD-free life expectancy of 31·46 [30·36-32·48] years, while those with healthy sleep had 33·26 [33·08-33·46] years (difference: 1·80 [0·96-2·75]). The Women’s intermediate group lost 0.48 [0·41-0·55] life years (**Table 2, Figure 3**). A similar pattern emerged among males, yet more pronounced than among women. Poorly sleeping men expected a 27·96 [26·80-29·02] CVD-free life years, and healthy sleepers had 30·27 [30·07-30·49] years (difference: 2·31 [1·46-3·29]). The composite sleep categories suggested joint effects of different sleep characteristics compared to models only focusing on a single characteristic (**Table 2, appendix pp 21**). The corresponding hazard ratios (HR) for CVD were 1·13 [1·05-1·22] and 1·17 [1·09-1·26] in poor sleepers compared to healthy ones among females and males (**appendix pp 22-25**).

**Table 2.**
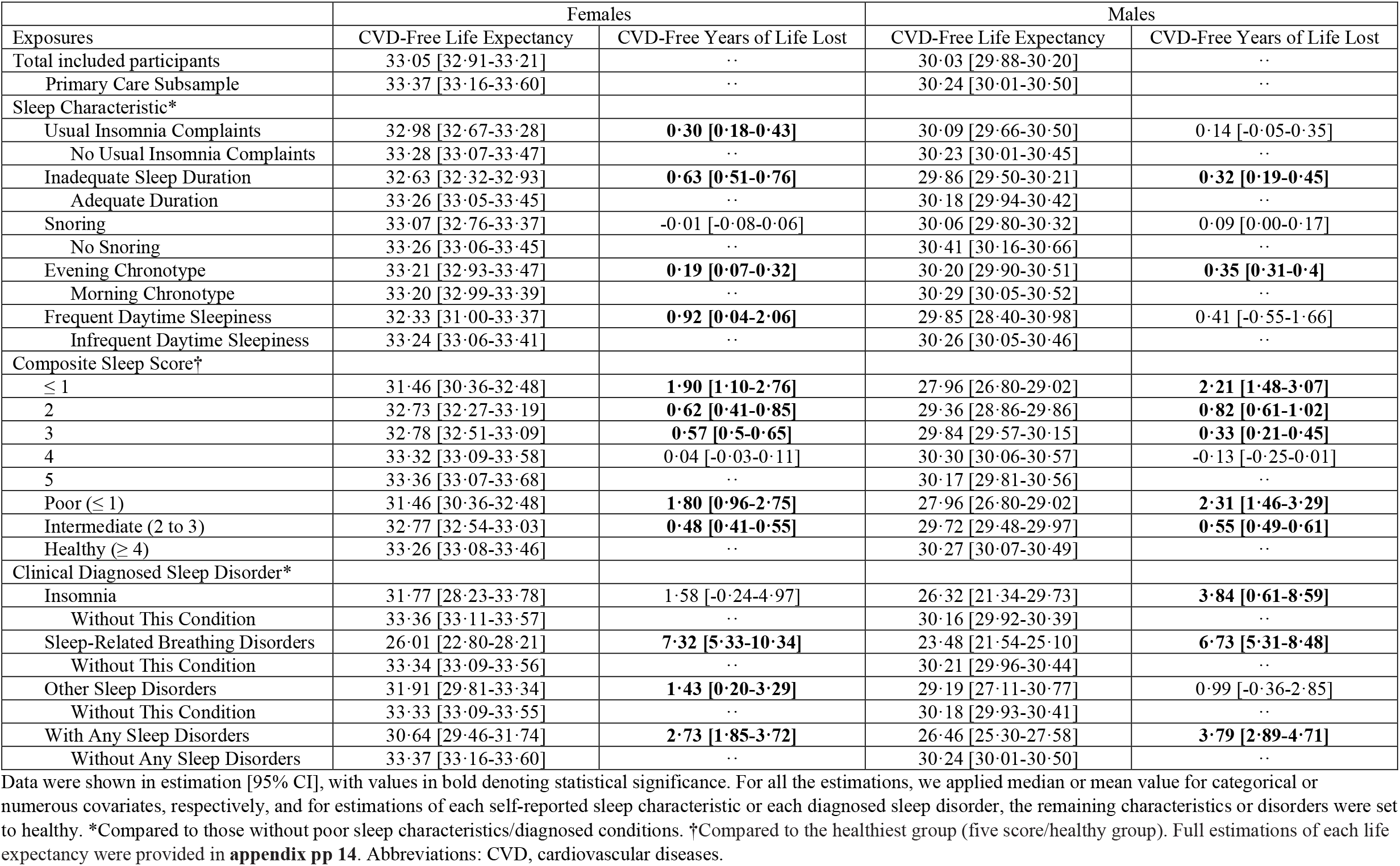
Cardiovascular-free life expectancy and years of life lost at age 40 among participants with different sleep exposures

**Figure 2.**
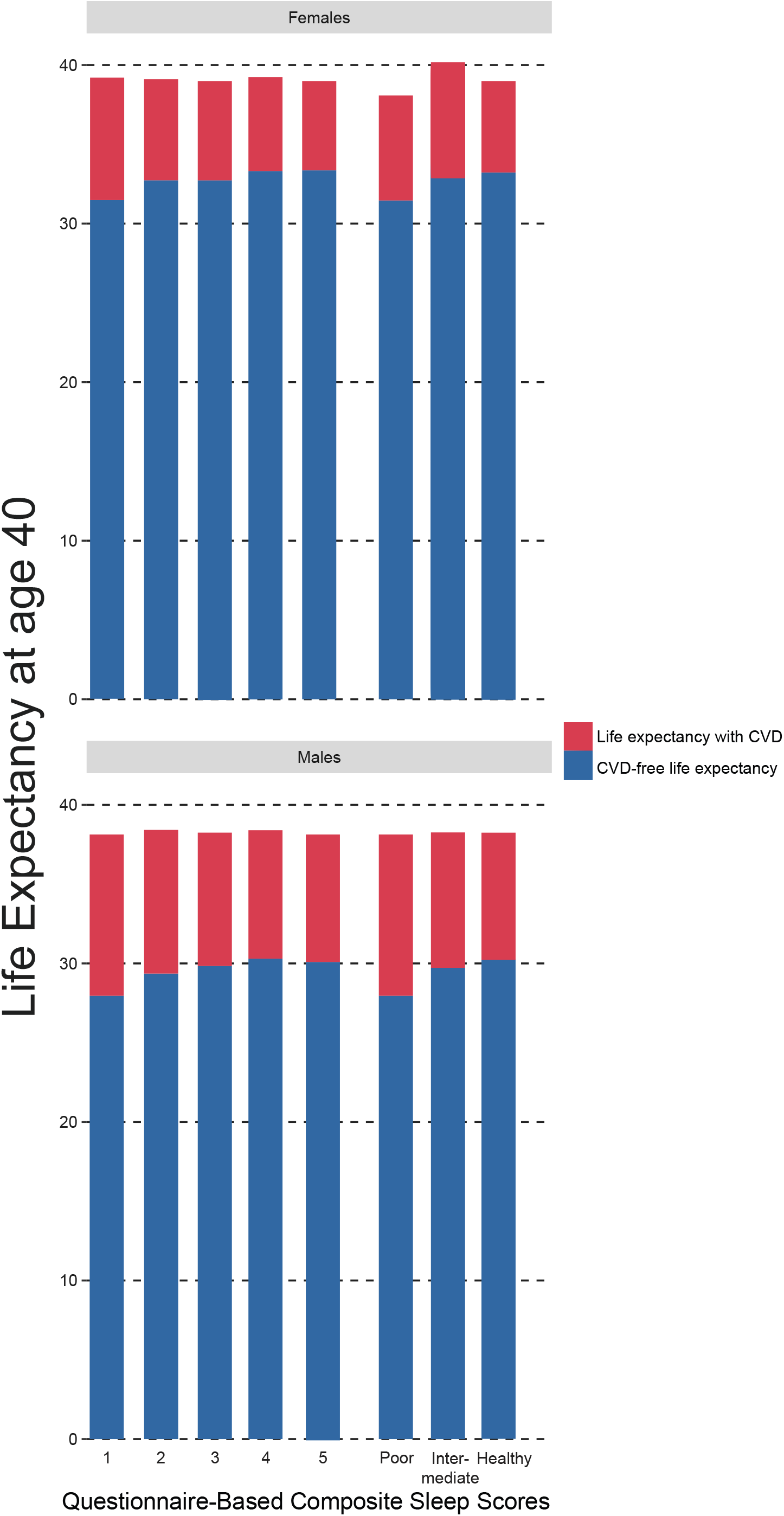
Total and cardiovascular disease-free life expectancy at age 40 among participants with different (A) questionnaire-based composite sleep scores or (B) diagnosed sleep disorders The results were adjusted for age, socioeconomic status, mental health issues, perceived health, body mass index (BMI), economic activity and shift work, cigarette smoking, alcohol consumption, diet quality, discretionary screen time, and physical activity. Full estimations of each life expectancy were provided in **appendix pp 14**.

Among the 140181 participants with primary care data available, 1203 had an insomnia diagnosis in the last two years before their enrollment, while 2441 and 6489 were diagnosed with sleep-related breathing disorders and other sleep disorders (**appendix pp14-15**). A confusion matrix confirmed an assured negative predictive value of self-reported with diagnoses (**appendix pp 18**). During an average follow-up of 11.8 (1.2) years and 1655597 person-years at risk of death, 5996 died, and 23480 developed CVD. Total life expectancy was an additional 39·22 [39·05-39·37] and 38·29 [38·09-38·47] years (**Figure 2B, appendix pp 16**), while CVD-free life expectancies were 33·37 [33·16-33·60] and 30·24 [30·01-30·50] years for females and males (**Table 2, Figure 2B**). Contrary to self-report sleep, participants with any clinical sleep disorders lost up to 1·16 [0·43-2·21] life compared to those without (**appendix pp 16**). Those diagnosed with sleep disorders showed similar characteristics to those with poor self-reported sleep (**appendix pp14-15**). Women diagnosed with sleep-related breathing disorders or other sleep disorders lost 7·32 [5·33-10·34] (HR: 2·09 [1·87-2·35]) or 1·43 [0·2-3·29] (HR: 1·20 [1·08-1·34]) CVD-free years of life compared to those without the corresponding diagnosis, respectively (**Table 2, Figure 3, appendix pp 26-29**). In men, those with insomnia or sleep-related breathing disorders expected a significant CVD-free life loss by 3·84 [0·61-8·59] (HR: 1·49 [1·23-1·80]) or 6·73 [5·31-8·48] (HR: 1·83 [1·68-1·99]) years, respectively. Complete estimations of life expectancy and HRs were provided in the **appendix pp 16, 22-29**.

**Fig. 3.**
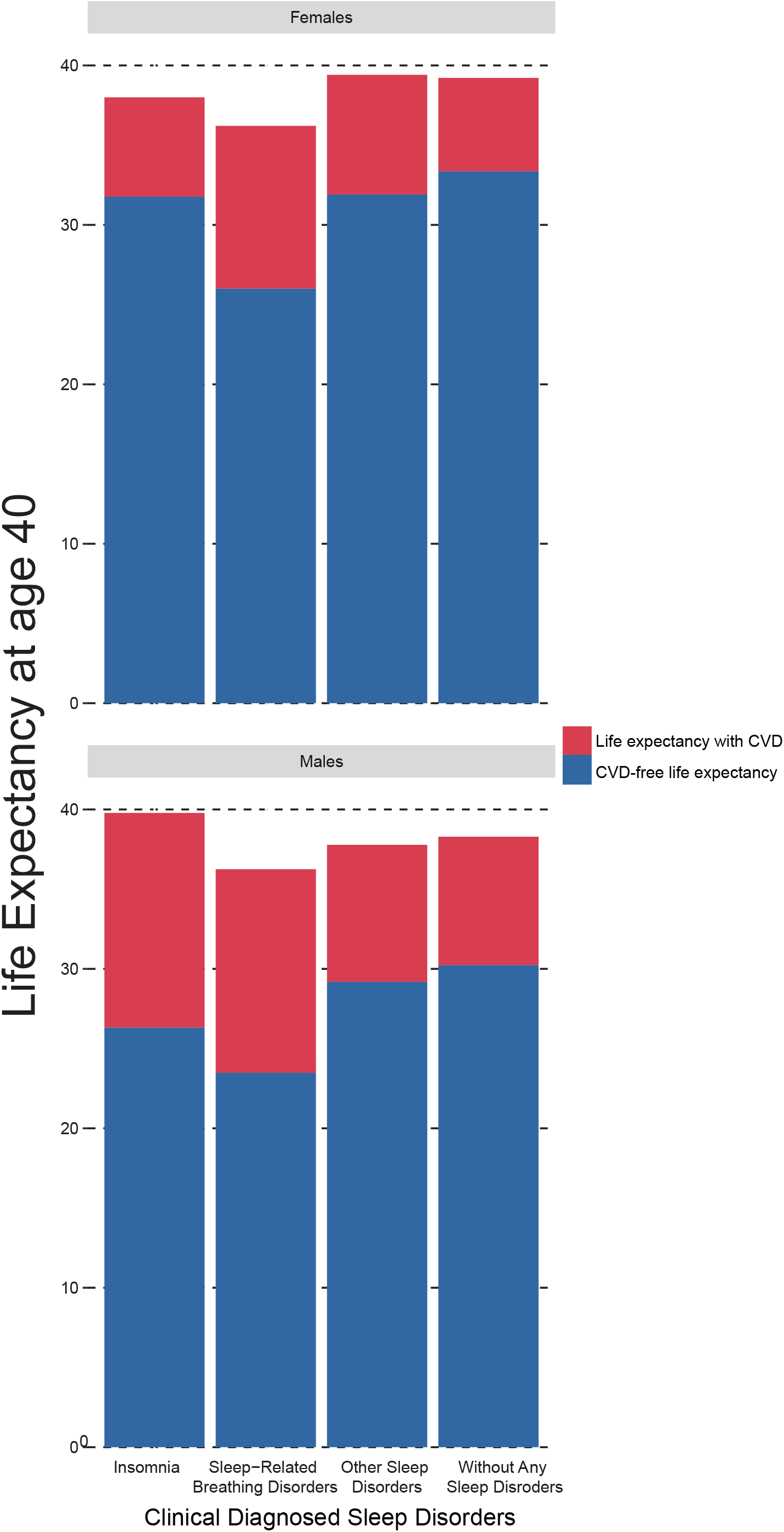

## Discussion

To our knowledge, the current study provides the first comprehensive analyses of sleep measures with life expectancy, utilizing cohort study data and primary care electronic health records to identify sleep disorders. At 40 years of age, participants with self-reported poor sleep lost 1·8 and 2·3 years of life free of CVD in women and men, respectively, compared to those who reported healthy sleep. The diagnosis of sleep-related breathing disorders was associated with the largest loss of life years without diseases in both sexes, up to 7·3 years, while diagnosed insomnia compromised life years in men by 3·8 years. Our findings strongly suggest that people with either self-reported or diagnosed unhealthy sleep have less life expectancy free of CVD.

“Seven to 9 hours of good-quality sleep on a regular basis, with consistent bed and wake-up times” is the only national (Canada) guideline on sleep,(3) based on a review of reviews on duration and a *de novo* systematic review of the regularity/consistency and chronotype/timing.(5,6) Although we could not address sleep regularity/consistency as it was outside the scope of the UK Biobank questionnaires, aligned with this guideline, our finding supported that sleep duration and chronotype impact CVD-free life expectancy in both sexes. However, “sleep quality” is a poorly defined term encompassing various measures and characteristics, so in the present study, we applied available CVD risk factors, including insomnia complaints, snoring, and daytime sleepiness. Despite the controversial outcomes between recent systematic reviews of insomnia,(25,26) our analyses showed that insomnia impacted CVD-free life expectancy differently between sexes, as highlighted by the distinct effects of self-reported complaints and clinical insomnia. Although we found no association of self-reported/co-sleeper-reported snoring with life expectancy outcomes, clinical sleep-related breathing disorders were found to compromise CVD-free life expectancy in both sexes by up to 7·3 years. Our results are consistent with those from the only existing review on topic ^26^ and suggest that women with frequent daytime sleepiness impacted the number of CVD-free years. The absence of clinical diagnosis of excessive daytime sleepiness and the low prevalence (*i*.*e*., 33 out of 140181 participants) of circadian rhythm sleep disorders, the corresponding clinical diagnosis of chronotype, prohibited us from further investigating objective measurements.

Besides the independent effect of each characteristic, composite sleep measures provide more comprehensive insights and a more coherent public health message(3) and are increasingly used in cohort studies.(8,9) Although there is still no systematic review available on composite sleep exposures, the present results, consistent with existing individual studies,(8,9) suggested that people with multiple poor sleep characteristics had a shorter CVD-free year in both sexes. Our study further suggested that this association is more prominent in males, while the very few longitudinal results of sleep and health examining sex as an effect modifier are incongruous.(6,27–29) The systematic review conducted by He et al. (2020) found that the mortality risk of long sleep duration was higher in females, but the mortality risk of short sleep was higher in males.(27) However, the systematic review of the Canadian national guideline and recent prospective studies found no sex differences in the mortality and CVD risks of non-adequate sleep duration.(6,28,29) Since capacities of sleep measurements vary between cohorts, arbitrariness in the included sleep measures and the subsequent adjustments hamper the aggregation of findings. More studies with detailed outcomes on each sleep measure are warranted to facilitate future harmonization and consensus on essential sleep characteristics selections.

This study’s main strength and novelty is the use of primary care data linkage to identify sleep disorders, allowing us to compare the differences between clinical sleep disorders and self-reported poor sleep with detailed sleep questionnaires. The large sample size further allows us to explore effect size with adequate statistical power. National hospitalization and death records across all participants reduce the potential measurement bias of outcomes and allow us to perform life expectancy analyses comprising morbidity and mortality. However, our study has several limitations. First, sleep-related clinical events other than insomnia and sleep-related breathing disorders were rare and thereby had to be grouped, compromising the specificity of our analyses. Second, the UK Biobank has poor representativeness by oversampling the high socioeconomic status, while our exclusion criteria further oversampled the healthy population (**Appendix pp 9-11**), so our findings might not be generalizable to a population with different socioeconomic status. The low prevalence of clinical sleep disorders also indicated a potential underestimation due to the undiagnosed. However, recent UK Biobank evidence suggests that poor representativeness is unlikely to influence the associations between lifestyle exposures and mortality.(30) Third, we could not rule out immortal-time bias as the current analyses only followed up mid-age healthy adults. Fourth, we could not distinguish CVD recovery and relapse events based on available data, so we could not avoid overestimating the time spent on diseases. Fifth, we only considered baseline measurements of exposures and covariates to maintain statistical power, although our previous finding suggested a reasonable temporal consistency.(8) Lastly, in the current study, we conservatively chose underlying conditions as covariables since comorbidities could be on the causal pathway (*i*.*e*., mediators). We could not eliminate the potential residual confounding (**Appendix pp 20**), so the true causality could be biased.

## Conclusions

In summary, this study provides further support for the advantage of a composite sleep score based on easily applied self-reported questions. With access to primary care records, our findings shed light on how clinical sleep disorders could be adversely associated with CVD-free life expectancy. These results could lay the cornerstone for future research on early prevention against poor sleep and improving CVD-free life expectancy among poor sleepers. Especially, we hope that the current outcomes can serve as a reference for policymakers and professionals to develop national sleep guidelines.

## Data Availability

The data that support the findings of this study are available from the UK Biobank but restrictions apply to the availability of these data, which were used under license for the current study, and so are not publicly available. Data are however available from the authors upon reasonable request and with permission of the UK Biobank.

https://www.ukbiobank.ac.uk/

## List of Abbreviations

BMI: Body Mass Index
BNF: British National Formulary
CTV3: Clinical Terms Version 3
CVD: Cardiovascular Disease
dm+d: Dictionary of Medicines and Devices
HR: Hazard Ratios
ICD: International Classification of Diseases
NHS: National Health Service
Read v2: Read codes version 2

## Declarations

### Ethics approval

This study was a secondary analysis of previously collected data so additional ethical approval was not required. The National Health Service and the National Research Ethics Service have approved the UK Biobank (Ref. 11/NW/0382) while all participants gave written informed consent.

### Consent for publication

Not applicable.

## Authors’ contributions

BH: conceptualization, software, formal analysis, visualization, writing – original draft. BPC & AT: methodology, software, validation, writing – review & editing. PC: conceptualization, methodology, writing – review & editing. ES: funding acquisition, project administration, resources, supervision, writing – review & editing.

## Acknowledgements

This research has been conducted using the UK Biobank Resource under Application Number 25813. The authors gratefully thank all the participants and professionals contributing to the UK Biobank. In addition, the authors acknowledge the Sydney Informatics Hub and the Artemis University of Sydney’s high-performance computing cluster Artemis for providing the computing resources and support to complete the research reported in this paper.

---

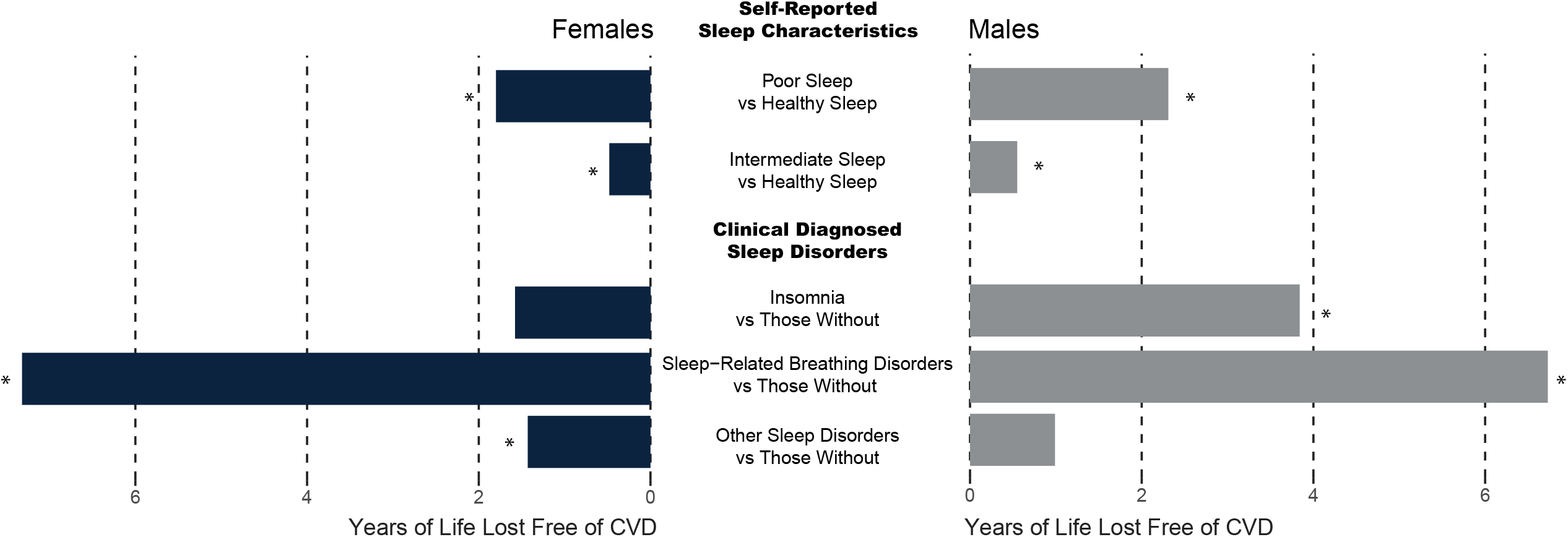

