## Supplementary Materials for "Influence of Poor Sleep on Cardiovascular Disease-Free Life Expectancy: A Multi-Resource-Based Population Cohort Study"

### Supplemental Material

**Supplemental Methods**

#### Supplemental Methods

##### Method S1. Primary Care Data Linkage, Cleaning, and Harmonization

The Phoenix Partnership (TPP) and the Egton Medical Information Systems Health (EMIS) supply primary care data for England and Wales/Scotland, respectively, while the Vision Health provides data across the UK.

Primary care clinical events are coded with Read codes version 2 (Read v2) in Scotland and Wales, or in both Read v2 and Clinical Terms Version 3 (CTV3) in England, depending on the data providers. Firstly, we converted both Read v2 and CTV3 into the following coding systems (ordered in priority), International Classification of Diseases (ICD) 10, Office of Population Censuses and Surveys Classification of Surgical Operations and Procedures (OPCS) 4, and ICD-9. Prescription codes shown in clinical events were excluded as misclassification. Then, the residual Read v2 records were all converted into CTV3. Finally, CTV3 was directly (look-up, LKP) referred to clinical events. As shown in **Fig. 1**., 7.4%, 2.1%, and 0.2% of records were recoded into ICD-10, OPSC-4, and ICD-9, respectively.

Primary care prescriptions are coded diversely. In Wales, only Read v2 was used. In Scotland, British National Formulary (BNF) and corresponding Read v2 were recorded, with some records only in Read v2. In England, TPP applies a modified version of BNF (TPPBNF) with a lower resolution compared to the original BNF, while Vision uses Dictionary of Medicines and Devices (dm+d), with some records with corresponding Read v2 available. As shown in **Fig. 2**., we tried to convert all records into BNF. In this study, 96.0% records were included with at least Section details available (see the BNF website for the definition of Section, https://openprescribing.net/bnf/).

##### Method S2. Detailed Start and Censor for Each Health-Related Data Linkage

Inpatient records started from 1997, 1998, and 1981, while censoring on 19 March 2021, 20 March 2021, and 15 Apr 2020, for England, Wales, and Scotland, respectively. Death records started from the UK Biobank enrolment and censored on 22 February 2021 and 03 March 2021 for England/Wales and Scotland, respectively. Primary care records started from the date when participants registered for a primary care service, as early as 1 January 1948 (England Vision), 5 July 1942 (England TPP), 9 November 1946 (Wales), or 31 December 1942 (Scotland), and censored on primary care resignment or data linkage censoring date (8 June 2017 (England Vision), 20 September 2018 (England TPP), 19 April 2017 (Wales), or 15 June 2016 (Scotland)) whichever applicable came first. The median dates when participants registered for a primary care service were 08 November 1999 (England Vision), 26 June 1997 (England TPP), 9 June 1988 (Wales), or 24 March 1993 (Scotland).

#### Supplemental Tables

**Table S1. Definitions of Cardiovascular Disease**

| Condition | ICD-9 | ICD-10 |
|---|---|---|
| Cardiovascular disease (CVD) (excluding essential (primary) hypertension, hypertensive renal disease, secondary hypertension, pulmonary embolism, other diseases of pulmonary vessels, other heart disorders in diseases classified elsewhere, oesophageal varices, varicose veins of other sites, but including senility) | 390 to 398, 402, 404, 411 to 415.0, 416, 417, 420 to 449, 451 to 455, 456.3 to 456.6, 456.8, 459 to 459, 797 | I0, I11, I13, I21 to I25, I27, I3, I4, I50, I51, I6, I7, I80 to I84, I87 to I9, R54 |

**Table S2. Original Questions for Self-Reported Sleep Characteristics**

| Characteristic | UK Biobank Code | UK Biobank Questionnaire | Healthy Answer | Unhealthy Answer |
|---|---|---|---|---|
| Insomnia Complaint | 1200 | Do you have trouble falling asleep at night or do you wake up in the middle of the night? | Never/rarely; Sometimes | Usually |
| Sleep Duration | 1160 | About how many hours sleep do you get in every 24 hours? (please include naps) (in integer). | 7 to <9 hr/d. | <7 or >=9 hr/d. |
| Snoring | 1210 | Does your partner or a close relative or friend complain about your snoring? | No | Yes |
| Chronotype | 1180 | Do you consider yourself to be? | Definitely a “morning” person; More a “morning” than “evening” person. | More an “evening” than a “morning person; Definitely an “evening” person. |
| Daytime Sleepiness | 1220 | How likely are you to doze off or fall asleep during the daytime when you don't mean to? (e.g. when working, reading or driving) | Never/rarely; Sometimes | Often; All the Time |

**Table S3. Definitions of Clinical Sleep Disorders in ICD and OPCS**

| Condition | ICD-9 | ICD-10/ OPCS4 |
|---|---|---|
| Insomnia | 327.0, 307.44, 780.61, 780.52 | G47.0, F51.0, Z72.820 |
| Hypersomnia | 327.1, 780.53, 780.54 | G47.1, G47.4, F51.1 |
| Sleep-related breathing disorders | 327.2, 780.57 | G47.3, E66.2, R06.3, R06.00, R06.8, R06.09 |
| Circadian rhythm sleep disorders | 327.3, 307.45, 780.45 | G47.2 |
| Parasomnias (including sleep-related bruxism) | 327.4, 307.42, 307.46, 307.47, 307.48, 307.49, 780.56, 780.59 | G47.5, F51.3, F51.4, F51.5, F51.8 |
| Non-specific | 327, 307.4, 780.5 | G47, F51 |

Abbreviations: ICD, International Classification of Diseases; OPCS, Office of Population Censuses and Surveys Classification of Surgical Operations and Procedures.

**Table S4. Definitions of Clinical Sleep Disorders in CTV3**

| Condition | CTV3 | LKP Description |
|---|---|---|
| Insomnia | .1B1B | Cannot sleep - insomnia |
| | .1BX0 | Delayed onset of sleep |
| | .E4A. | Insomnia |
| | .R051 | [D]Insomnia with sleep apnoea |
| | .R052 | [D]Insomnia |
| | 1B1B. | C/O - insomnia |
| | 1B1B0 | Initial insomnia |
| | 1B1B1 | Middle insomnia |
| | 1B1B2 | Late insomnia |
| | 1BX0. | Delayed onset of sleep |
| | E2741 | Insomnia NOS |
| | E2742 | Persistent insomnia |
| | Eu510 | Nonorganic insomnia |
| | Fy00. | Disorders of initiating and maintaining sleep |
| | R0051 | [D]Insomnia with sleep apnoea |
| | R0052 | [D]Insomnia NOS |
| | X007s | Insomnia NOS |
| | X007t | Nonorganic insomnia |
| | X007u | Cannot get off to sleep |
| | X0089 | Delayed sleep phase syndrome |
| | X76AF | Cannot sleep at all |
| | Xa7wV | Difficulty sleeping |
| | XaIti | Delayed onset of sleep |
| | XE0ux | Cannot sleep - insomnia |
| | XE2Pv | Insomnia |
| | XM0CT | C/O - insomnia |
| | XM0yu | [D]Insomnia |
| Hypersomnia | .1BX1 | Excessive sleep |
| | .R053 | [D]Hypersomnia with sleep apnoea |
| | .R054 | [D]Hypersomnia |
| | 1BX1. | Excessive sleep |
| | E274. | Hypersomnia of non-organic origin |
| | E2743 | Hypersomnia NOS |
| | E2744 | Persistent hypersomnia |
| | Eu511 | Hypersomnia of non-organic origin |
| | Fy06. | Kleine-Levin syndrome |
| | R0054 | [D]Hypersomnia NOS |
| | X007w | Episodic cluster headache |
| | X007x | Hypersomnia NOS |
| | X007y | Hypersomnia of non-organic origin |
| | X008E | Kleine-Levin syndrome |
| | X76AQ | Cannot wake up |

| | | |
|---|---|---|
| | XaC0p | [D]Hypersomnia |
| | XE2nU | [D]Hypersomnia with sleep apnoea |
| Sleep-related breathing disorders | .F36Z | Ondine's curse |
| | .H66. | SAS - Sleep apnoea syndrome |
| | .R053 | [D]Hypersomnia with sleep apnoea |
| | F28yz | Ondine's curse |
| | Fy03. | Obstructive sleep apnoea |
| | Fy04. | Obstructive sleep apnoea |
| | H5B.. | SAS - Sleep apnoea syndrome |
| | H5B0. | Obstructive sleep apnoea |
| | Q318. | Ondine's curse |
| | X0083 | SAS - Sleep apnoea syndrome |
| | X0084 | Obstructive sleep apnoea |
| | X0085 | CSA - Central sleep apnoea |
| | X0086 | Mixed sleep apnoea |
| | X0087 | Alveolar sleep apnoea |
| | X008F | Ondine syndrome, Sleep-related respiratory failure |
| | X00pU | Pharyngeal operation for obstructive sleep apnoea and snoring |
| | X20M9 | Insertion of appliance for sleep apnoea |
| | Xa08C | Ondine's curse |
| | XaEGP | [D]Sleep apnoea syndrome |
| | XE18D | Ondine's curse |
| | XE2nU | [D]Hypersomnia with sleep apnoea |
| | XM0Go | Appliance for sleep apnoea |
| Circadian rhythm sleep disorders | .1BX3 | Early morning waking |
| | .E4A1 | Early waking |
| | .R055 | [D]Sleep rhythm inversion |
| | .R056 | [D]Sleep rhythm irregular |
| | .R057 | [D]Sleep-wake rhythm non-24-hour cycle |
| | 1BX3. | Early morning waking |
| | E274F | Inversion of sleep rhythm |
| | Eu512 | [X] Nonorganic disorder of the sleep-wake schedule: [psychogenic inversion of circadian rhythm (including nyctohemeral rhythm and sleep rhythm)] |
| | Fy02. | Circadian dysregulation |
| | R0055 | [D]Sleep rhythm inversion |
| | R0056 | [D]Sleep rhythm irregular |
| | R0057 | [D]Sleep-wake rhythm non-24-hour cycle |
| | X0080 | 24 hour hypersomnolence |
| | X0082 | Excess daytime sleepiness with sleep paralysis |
| | X008A | Non-24 hour sleep-wake cycle |
| | X76AE | Sleep rhythm problem |
| | X76AK | Early waking |
| | XaIv5 | Early morning waking |
| | XE1Zr | Non-organic disorder of the sleep-wake schedule |
| | XM06j | Irregular sleep-wake pattern |
| | XM06k | Circadian dysregulation |
| Parasomnias (including sleep-related bruxism) | .1B1D | C/O nightmares |
| | .663N | Asthma disturbing sleep |
| | .66Yg | Chronic obstructive pulmonary disease disturbs sleep |
| | .E4A0 | Night terrors |
| | .E4A2 | Nocturnal sleep-related eating disorder |
| | .R058 | [D]Sleep dysfunction with sleep stage disturbance |
| | .R059 | [D]Sleep dysfunction with arousal disturbance |
| | .R05Z | [D]Sleep dysfunction NEC |
| | 1B1D. | C/O nightmares |
| | 663N. | Asthma disturbing sleep |

| | | |
|---|---|---|
| | 663N0 | Asthma causing night waking |
| | 663N1 | Asthma disturbs sleep weekly |
| | 663N2 | Asthma disturbs sleep frequently |
| | 66Yg. | Chronic obstructive pulmonary disease disturbs sleep |
| | A86.. | Sleeping sickness |
| | E2747 | Sleepwalking |
| | E2748 | Night terrors |
| | E2749 | Nightmares |
| | E274A | Sleep drunkenness |
| | E274B | Repeated rapid eye movement sleep interruptions |
| | E274C | Other sleep stage or arousal dysfunction |
| | E274D | (Repetitive intrusions of sleep) or (restless sleep) |
| | E274y | (Dreams) or (other non-organic sleep disorder) |
| | Eu513 | Sleepwalking |
| | Eu514 | Night terrors |
| | Eu515 | [X] Nightmares or dream anxiety disorder |
| | Eu51y | [X]Other nonorganic sleep disorders |
| | Fy01. | Disorders of excessive somnolence |
| | Fy05. | Nocturnal sleep-related eating disorder |
| | Fyu58 | [X]Other sleep disorders |
| | R0058 | [D]Sleep dysfunction with sleep stage disturbance |
| | R0059 | [D]Sleep dysfunction with arousal disturbance |
| | R005z | [D]Sleep dysfunction NOS |
| | X008H | Parasomnia |
| | X764D | Low level of awareness whilst sleep walking |
| | X764E | Low level of reactivity whilst sleep walking |
| | X764F | Low level of motor skill whilst sleep walking |
| | X764G | Blank, staring face whilst sleep walking |
| | X764H | Unresponsive to communication whilst sleep walking |
| | X764I | Not easily wakened from sleep walking |
| | X764J | No recollection of sleep walk |
| | X764K | Sleep automatism |
| | X76AJ | Wakes and cannot sleep again |
| | X76AL | Circumstances interfere with sleep |
| | X76AM | Symptoms interfere with sleep |
| | XaKv8 | Chronic obstructive pulmonary disease disturbs sleep |
| | XE1Yi | Repetitive intrusions of sleep |
| | XM06i | Dyssomnia |
| Non-specific | .1B1Q | Poor sleep pattern |
| | .1BX9 | Light sleep |
| | .E4A. | Sleep disorders (& [insomnia] or [nightmares] or [sleepwalking (& somnambulism)]) |
| | .R05. | [D]Sleep disturbances (& [hypersomnia] or [insomnia]) |
| | .R050 | [D]Sleep disturbance, unspecified |
| | 1B1Q. | Poor sleep pattern |
| | 1BX9. | Light sleep |
| | E274. | Non-organic sleep disorders (& [hypersomnia] or [insomnia]) |
| | E2740 | Unspecified non-organic sleep disorder |
| | E274z | Non-organic sleep disorder NOS |
| | Eu51. | Non-organic sleep disorder |
| | Eu51z | [X]Nonorganic sleep disorder, unspecified |
| | Fy0.. | Sleep disorders |
| | R005. | [D]Sleep disturbances |
| | R0050 | [D]Sleep disturbance, unspecified |
| | R0053 | ([D]Hypersomnia with sleep apnoea) or (sleep apnoea syndrome) |
| | Ua0Lq | Sleeping rough |

| | | |
|---|---|---|
| | Ua1FL | Disturbing sleep |
| | X007q | Sleep disorders |
| | X007v | Broken sleep |
| | X76AG | Not getting enough sleep |
| | X76AN | Restless sleep |
| | X76AO | Unrefreshed by sleep |
| | Xa7wV | Difficulty sleeping |
| | XaFqr | Poor sleep pattern |
| | XE1bI | Sleep disorders (& [insomnia] or [nightmares] or [sleepwalking (& somnambulism)]) |
| | XE1gP | [D]Sleep dysfunction NEC |
| | XE2cd | [D]Sleep disturbances (& [hypersomnia] or [insomnia]) |
| | XE2Q5 | Non-organic sleep disorder |
| | XSGLz | Light sleep |

Abbreviations: CTV, Clinical Terms Version; LKP, directly LooK-Up.

**Table S5. Definition of Sleep Pill Prescription in BNF.**

| **BNF Description** | **BNF Code** |
|---|---|
| Chapter 4: Central Nervous System | 04 |
| Section 1: Hypnotics and anxiolytics | 0401 (The definition used in the current study) |
| Paragraph (subsection) 1: Hypnotics | 040101 |
| Paragraph (subsection) 2: Anxiolytics | 040102 |
| Paragraph (subsection) 3: Barbiturates | 040103 |

Abbreviations: BNF, British National Formulary.

**Table S6. Original Questions and Definitions for Covariates**

| Variable | UK Biobank Code | UK Biobank Questionnaire/Definition | Modification in the Present Study | Category |
|---|---|---|---|---|
| Socioeconomic status (Townsend deprivation index) | 189 | Each participant is assigned a score corresponding to the output area in which their postcode is located. The higher the score, the more deprived the area. | Derived into quantiles. | 1st quantile;<br>2nd quantile;<br>3rd quantile;<br>4th quantile. |
| Mental health issue | 2090, 2100 | Have you ever seen a general practitioner (GP)/ psychiatrist for nerves, anxiety, tension or depression? | If either is true then true. | Yes;<br>No |
| Perceived health | 2178 | In general how would you rate your overall health? | NA | Excellent; Good;<br>Fair; Poor |
| Body mass index (BMI) | 21001 | BMI value here is constructed from height and weight measured during the initial Assessment Centre visit. | Derived into three groups. | <30;<br>30 to <40;<br>>= 40 |
| Economic activity and shift work | 6142, 3426, 826 | Which of the following describes your current situation? (In paid employment or self-employed; Retired; Looking after home and/or family; Unable to work because of sickness or disability; Unemployed; Doing unpaid or voluntary work; Full or part-time student)<br>Does your work involve night shifts/ shift work? (Never/rarely; Sometimes; Usually; Always) | Four derived groups based on 'Current employment status' (6142), 'Job involves night shift work' (3426), and 'Job involves shift work' (826). | retired/not in the workforce;<br>employed not in shift work;<br>employed in night shift work;<br>employed in day shift work |
| Cigarette smoking | 20116 | This field summarizes the current/past smoking status of the participant. | NA | Never;<br>Previous smoker; Current smoker |
| Alcohol consumption | 1558, 1568, 1578, 1588, 1598, 1608, 5364.<br>20117 | The level of overall alcohol consumption as the number of UK units of alcohol (10 mL/unit) consumed per week was calculated; participants were categorized based on the consumption according to the UK guideline (14 UK units/wk). See https://biobank.ndph.ox.ac.uk/showcase/label.cgi?id=100051 for full questionnaire details. | | Never;<br>Previous drinker;<br>Within guidelines (14 units/wk) (including occasional drinkers);<br>Above guidelines (14-28 units/wk);<br>Double guidelines (>28 units/wk). |
| Diet quality | 1289, 1299, 1309, 1319, 1329, 1339, 1349, 1369, 1379, 1389, | Four healthy diet patterns were evaluated based on the American Heart Asociation (AHA), >4.5 servings/d fruits and vegetables, >= 2 times/wk fish intake, <2 times/wk processed meat, <5 times/wk red meat intake. Participants got one score for each criterion. We then categorized them based on the score. See https://biobank.ndph.ox.ac.uk/showcase/label.cgi?id=100052 for full questionnaire details. | | Poor (<= 1 score);<br>Intermediate (2 to 3 score);<br>Healthy (> 4 score). |
| Discretionary screen time | 1070, 1080 | In a typical DAY, how many hours do you spend watching TV/ the computer? (in integer) | The total daily hours of both. | (Continuous) hr/d |
| Physical activity | 864, 874, 884, 894, 904, 914 | The UK Biobank applied a the modified-version short-form International Physical Activity Questionnaire (IPAQ). We summarized weekly PA using weekly total Metabolic Equivalent Task (MET), calculated by multiplying the MET value of activity by the number of PA hours per week. Based on the lower and upper limits of the World Health Organization (WHO) PA guideline, we categorized PA into three groups. Another category "No moderate-to-vigorous PA" was further defined. See https://biobank.ndph.ox.ac.uk/showcase/label.cgi?id=100054 for full questionnaire details. | | No moderate-to-vigorous PA;<br>Low (0 to < 600 MET-mins/wk);<br>Medium (600 to < 1200 MET-mins/wk);<br>High (≥ 1200 MET-mins/wk). |

**Table S7. Comparison Sample Characteristics between Participants Excluded and Included**

| | | | | Excluded | | | Included | | |
|---|---|---|---|---|---|---|---|---|---|
| label | Total N | Missing N | Levels | Females | Males | Total | Females | Males | Total |
| Total N (%) | ·· | ·· | ·· | 99807 (52) | 93969 (48) | 193776 | 173546 (56) | 135137 (44) | 308683 |
| Chronotype | 135461 | 58315 | Evening | 27507 (28) | 23607 (25) | 51114 (26) | 63635 (37) | 51677 (38) | 115312 (37) |
| | | | Early | 46493 (47) | 37854 (40) | 84347 (44) | 109911 (63) | 83460 (62) | 193371 (63) |
| | | | (Missing) | 25807 (26) | 32508 (35) | 58315 (30) | ·· | ·· | ·· |
| Duration | 189562 | 4214 | Inadequate | 35659 (36) | 33166 (35) | 68825 (36) | 51765 (30) | 40992 (30) | 92757 (30) |
| | | | Adequate | 61586 (62) | 59151 (63) | 120737 (62) | 121781 (70) | 94145 (70) | 215926 (70) |
| | | | (Missing) | 2562 (3) | 1652 (2) | 4214 (2) | ·· | ·· | ·· |
| Insomnia | 192273 | 1503 | Usual | 35212 (35) | 25036 (27) | 60248 (31) | 51880 (30) | 29248 (22) | 81128 (26) |
| | | | No Usual | 63921 (64) | 68104 (72) | 132025 (68) | 121666 (70) | 105889 (78) | 227555 (74) |
| | | | (Missing) | 674 (1) | 829 (1) | 1503 (1) | ·· | ·· | ·· |
| Snore | 156726 | 37050 | Yes | 23003 (23) | 37598 (40) | 60601 (31) | 47873 (28) | 64873 (48) | 112746 (37) |
| | | | No | 53677 (54) | 42448 (45) | 96125 (50) | 125673 (72) | 70264 (52) | 195937 (63) |
| | | | (Missing) | 23127 (23) | 13923 (15) | 37050 (19) | ·· | ·· | ·· |
| Daytime Sleepiness | 190042 | 3734 | Frequent | 3321 (3) | 3689 (4) | 7010 (4) | 3692 (2) | 3391 (3) | 7083 (2) |
| | | | Not Frequent | 94682 (95) | 88350 (94) | 183032 (94) | 169854 (98) | 131746 (97) | 301600 (98) |
| | | | (Missing) | 1804 (2) | 1930 (2) | 3734 (2) | ·· | ·· | ·· |
| Follow-up Time (yr) | 193776 | 0 | Mean (SD) | 11·7 (1·9) | 11·4 (2·3) | 11·5 (2·1) | 11·9 (1·2) | 11·8 (1·4) | 11·8 (1·3) |
| Incident CVD | 193776 | 0 | No | 54271 (54) | 40031 (43) | 94302 (49) | 148595 (86) | 107024 (79) | 255619 (83) |
| | | | Yes | 19610 (20) | 22067 (23) | 41677 (22) | 24951 (14) | 28113 (21) | 53064 (17) |
| | | | Before Enrolment | 25926 (26) | 31871 (34) | 57797 (30) | ·· | ·· | ·· |
| All-Cause Death | 193776 | 0 | No | 91780 (92) | 81108 (86) | 172888 (89) | 167494 (97) | 127399 (94) | 294893 (96) |
| | | | Yes | 8027 (8) | 12861 (14) | 20888 (11) | 6052 (3) | 7738 (6) | 13790 (4) |
| Age of Enrollment | 193776 | 0 | Mean (SD) | 58·2 (7·8) | 58·7 (8·0) | 58·4 (7·9) | 56·1 (8·0) | 56·2 (8·2) | 56·2 (8·1) |
| Socioeconomic Status | 193153 | 623 | Highest Deprivation | 29192 (29) | 28582 (30) | 57774 (30) | 37778 (22) | 29908 (22) | 67686 (22) |
| | | | 2nd Deprivation | 25169 (25) | 22954 (24) | 48123 (25) | 44057 (25) | 33282 (25) | 77339 (25) |
| | | | 3rd Deprivation | 23396 (23) | 21549 (23) | 44945 (23) | 45431 (26) | 35130 (26) | 80561 (26) |
| | | | Least Deprivation | 21723 (22) | 20588 (22) | 42311 (22) | 46280 (27) | 36817 (27) | 83097 (27) |
| | | | (Missing) | 327 (0) | 296 (0) | 623 (0) | ·· | ·· | ·· |
| Mental Health Issues | 191599 | 2177 | No | 55008 (55) | 65928 (70) | 120936 (62) | 104680 (60) | 101522 (75) | 206202 (67) |

| | | | | | | | | | |
|---|---|---|---|---|---|---|---|---|---|
| | | | Yes | 43733 (44) | 26930 (29) | 70663 (36) | 68866 (40) | 33615 (25) | 102481 (33) |
| | | | (Missing) | 1066 (1) | 1111 (1) | 2177 (1) | ·· | ·· | ·· |
| Perceived Health | 190290 | 3486 | Excellent | 12021 (12) | 10453 (11) | 22474 (12) | 34298 (20) | 25079 (19) | 59377 (19) |
| | | | Good | 54607 (55) | 47079 (50) | 101686 (52) | 106933 (62) | 80374 (59) | 187307 (61) |
| | | | Fair | 24825 (25) | 26556 (28) | 51381 (27) | 28089 (16) | 25887 (19) | 53976 (17) |
| | | | Poor | 6540 (7) | 8209 (9) | 14749 (8) | 4226 (2) | 3797 (3) | 8023 (3) |
| | | | (Missing) | 1814 (2) | 1672 (2) | 3486 (2) | ·· | ·· | ·· |
| Body Mass Index (kg/m2) | 190672 | 3104 | <30 | 70879 (71) | 65742 (70) | 136621 (71) | 136713 (79) | 103783 (77) | 240496 (78) |
| | | | 30 to <40 | 24273 (24) | 24981 (27) | 49254 (25) | 33384 (19) | 29897 (22) | 63281 (21) |
| | | | >= 40 | 3196 (3) | 1601 (2) | 4797 (2) | 3449 (2) | 1457 (1) | 4906 (2) |
| | | | (Missing) | 1459 (1) | 1645 (2) | 3104 (2) | ·· | ·· | ·· |
| Economic Activity and Shift Work | 188025 | 5751 | Retired/not in the workforce | 50755 (51) | 44396 (47) | 95151 (49) | 70322 (41) | 44118 (33) | 114440 (37) |
| | | | Employed not in shift work | 37769 (38) | 36119 (38) | 73888 (38) | 89084 (51) | 74480 (55) | 163564 (53) |
| | | | Employed in night shift work | 3625 (4) | 6357 (7) | 9982 (5) | 5974 (3) | 9407 (7) | 15381 (5) |
| | | | Employed in day shift work | 4560 (5) | 4444 (5) | 9004 (5) | 8166 (5) | 7132 (5) | 15298 (5) |
| | | | (Missing) | 3098 (3) | 2653 (3) | 5751 (3) | ·· | ·· | ·· |
| Cigarette Smoking | 190828 | 2948 | Never | 57036 (57) | 41939 (45) | 98975 (51) | 105000 (61) | 69521 (51) | 174521 (57) |
| | | | Cessation | 31005 (31) | 37785 (40) | 68790 (35) | 54436 (31) | 49818 (37) | 104254 (34) |
| | | | Current | 10254 (10) | 12809 (14) | 23063 (12) | 14110 (8) | 15798 (12) | 29908 (10) |
| | | | (Missing) | 1512 (2) | 1436 (2) | 2948 (2) | ·· | ·· | ·· |
| Alcohol Drinking | 192121 | 1655 | Never | 7746 (8) | 3437 (4) | 11183 (6) | 8232 (5) | 2969 (2) | 11201 (4) |
| | | | Quit | 4695 (5) | 4283 (5) | 8978 (5) | 5285 (3) | 3836 (3) | 9121 (3) |
| | | | < 14 UK Units/wk | 67030 (67) | 40949 (44) | 107979 (56) | 116520 (67) | 56458 (42) | 172978 (56) |
| | | | 14 to <28 UK Units/wk | 14313 (14) | 23127 (25) | 37440 (19) | 31948 (18) | 37148 (27) | 69096 (22) |
| | | | >= 28 UK Units/wk | 5195 (5) | 21346 (23) | 26541 (14) | 11561 (7) | 34726 (26) | 46287 (15) |
| | | | (Missing) | 828 (1) | 827 (1) | 1655 (1) | ·· | ·· | ·· |
| Dietary Quality | 188965 | 4811 | Poor | 3964 (4) | 10276 (11) | 14240 (7) | 5795 (3) | 14142 (10) | 19937 (6) |
| | | | Intermediate | 57803 (58) | 59263 (63) | 117066 (60) | 100078 (58) | 88933 (66) | 189011 (61) |
| | | | Healthy | 36002 (36) | 21657 (23) | 57659 (30) | 67673 (39) | 32062 (24) | 99735 (32) |
| | | | (Missing) | 2038 (2) | 2773 (3) | 4811 (2) | ·· | ·· | ·· |
| Discretionary Screen Time (hr/d) | 188046 | 5730 | Mean (SD) | 3·8 (2·1) | 4·2 (2·4) | 4·0 (2·3) | 3·6 (1·9) | 4·0 (2·1) | 3·8 (2·0) |
| Physical Activity | 146652 | 47124 | No MVPA | 16526 (17) | 15802 (17) | 32328 (17) | 28252 (16) | 18958 (14) | 47210 (15) |
| | | | < 10 MET hrs/wk | 7275 (7) | 7689 (8) | 14964 (8) | 18041 (10) | 14267 (11) | 32308 (10) |

| | | | | | | | | | |
|---|---|---|---|---|---|---|---|---|---|
| | | | 10 to <20 MET hrs/wk | 10296 (10) | 10290 (11) | 20586 (11) | 27516 (16) | 19885 (15) | 47401 (15) |
| | | | >= 20 MET hrs/wk | 36879 (37) | 41895 (45) | 78774 (41) | 99737 (57) | 82027 (61) | 181764 (59) |
| | | | (Missing) | 28831 (29) | 18293 (19) | 47124 (24) | .. | .. | .. |

Data were shown in N (% vertically) or Mean (S.D.) for categorical or continuous variables, respectively. Abbreviations: CVD, cardiovascular diseases; MET, metabolic equivalent task; MVPA, moderate-to-vigorous physical activity

**Table S8. Basic Characteristics of Participants with Different Self-Reported Sleep Characteristics**

| Variables | Poor | Intermediate | Healthy | Total |
|---|---|---|---|---|
| n | 6479 (2) | 117374 (38) | 184830 (60) | 308683 |
| Early Chronotype | 516 (8) | 47763 (41) | 145092 (79) | 193371 (63) |
| Adequate Duration (7 to <9 hr/d) | 249 (4) | 50726 (43) | 164951 (89) | 215926 (70) |
| No Usual Insomnia | 299 (5) | 58972 (50) | 168284 (91) | 227555 (74) |
| No Snoring | 372 (6) | 49417 (42) | 146148 (79) | 195937 (63) |
| No Frequent Daytime Sleepiness | 4653 (72) | 112863 (96) | 184084 (100) | 301600 (98) |
| Follow-up Time (yr) | 11·8 (1·4) | 11·8 (1·3) | 11·9 (1·2) | 11·8 (1·3) |
| Incident CVD | 1568 (24) | 22134 (19) | 29362 (16) | 53064 (17) |
| All-Cause Death | 393 (6) | 5776 (5) | 7621 (4) | 13790 (4) |
| Age of Enrollment | 56·2 (7·7) | 56·4 (7·9) | 56·0 (8·2) | 56·2 (8·1) |
| Sex | | | | |
| Female | 3407 (53) | 61966 (53) | 108173 (59) | 173546 (56) |
| Male | 3072 (47) | 55408 (47) | 76657 (41) | 135137 (44) |
| Socioeconomic Status | | | | |
| Highest Deprivation | 1987 (31) | 27688 (24) | 38011 (21) | 67686 (22) |
| 2nd Deprivation | 1648 (25) | 29529 (25) | 46162 (25) | 77339 (25) |
| 3rd Deprivation | 1464 (23) | 29891 (25) | 49206 (27) | 80561 (26) |
| Least Deprivation | 1380 (21) | 30266 (26) | 51451 (28) | 83097 (27) |
| Mental Health Issues | 3286 (51) | 44125 (38) | 55070 (30) | 102481 (33) |
| Perceived Health | | | | |
| Excellent | 483 (7) | 16855 (14) | 42039 (23) | 59377 (19) |
| Good | 2893 (45) | 69029 (59) | 115385 (62) | 187307 (61) |
| Fair | 2201 (34) | 26763 (23) | 25012 (14) | 53976 (17) |
| Poor | 902 (14) | 4727 (4) | 2394 (1) | 8023 (3) |
| Body Mass Index (kg/m2) | | | | |
| <30 | 3863 (60) | 85456 (73) | 151177 (82) | 240496 (78) |
| 30 to <40 | 2274 (35) | 29307 (25) | 31700 (17) | 63281 (21) |
| >= 40 | 342 (5) | 2611 (2) | 1953 (1) | 4906 (2) |
| Economic Activity and Shift Work | | | | |
| Retired/not in the workforce | 2831 (44) | 44725 (38) | 66884 (36) | 114440 (37) |
| Employed not in shift work | 2798 (43) | 59436 (51) | 101330 (55) | 163564 (53) |
| Employed in night shift work | 491 (8) | 7078 (6) | 7812 (4) | 15381 (5) |
| Employed in day shift work | 359 (6) | 6135 (5) | 8804 (5) | 15298 (5) |
| Cigarette Smoking | | | | |
| Never | 2937 (45) | 61107 (52) | 110477 (60) | 174521 (57) |
| Cessation | 2467 (38) | 42017 (36) | 59770 (32) | 104254 (34) |
| Current | 1075 (17) | 14250 (12) | 14583 (8) | 29908 (10) |
| Alcohol Drinking | | | | |
| Never | 234 (4) | 4006 (3) | 6961 (4) | 11201 (4) |
| Quit | 312 (5) | 3581 (3) | 5228 (3) | 9121 (3) |
| < 14 UK Units/wk | 3353 (52) | 62239 (53) | 107386 (58) | 172978 (56) |
| 14 to <28 UK Units/wk | 1240 (19) | 26460 (23) | 41396 (22) | 69096 (22) |
| >= 28 UK Units/wk | 1340 (21) | 21088 (18) | 23859 (13) | 46287 (15) |

| | | | | |
|---|---|---|---|---|
| Dietary Quality | | | | |
| Poor | 717 (11) | 9456 (8) | 9764 (5) | 19937 (6) |
| Intermediate | 4123 (64) | 73499 (63) | 111389 (60) | 189011 (61) |
| Healthy | 1639 (25) | 34419 (29) | 63677 (34) | 99735 (32) |
| Discretionary Screen Time (hr/d) | 4·6 (2·6) | 4·0 (2·1) | 3·6 (1·9) | 3·8 (2·0) |
| Physical Activity | | | | |
| No MVPA | 1579 (24) | 20628 (18) | 25003 (14) | 47210 (15) |
| < 10 MET hrs/wk | 852 (13) | 13662 (12) | 17794 (10) | 32308 (10) |
| 10 to <20 MET hrs/wk | 896 (14) | 18093 (15) | 28412 (15) | 47401 (15) |
| >= 20 MET hrs/wk | 3152 (49) | 64991 (55) | 113621 (61) | 181764 (59) |

Data were shown in N (% vertically) or Mean (S.D.) for categorical or continuous variables, respectively.

**Table S9. Basic Characteristics of Participants with Different Diagnosed Sleep Disorders**

| | Females | | | Males | | | All | | |
|---|---|---|---|---|---|---|---|---|---|
| Variables | None | With Any Disorders | Total | None | With Any Disorders | Total | None | With Any Disorders | Total |
| n | 75564 (95) | 3662 (5) | 79226 | 58030 (95) | 2925 (5) | 60955 | 133594 (95) | 6587 (5) | 140181 |
| Insomnia | ·· | 853 (23) | 853 (1) | ·· | 350 (12) | 350 (1) | ·· | 1203 (18) | 1203 (1) |
| Sleep-Related Breathing Disorders | ·· | 957 (26) | 957 (1) | ·· | 1484 (51) | 1484 (2) | ·· | 2441 (37) | 2441 (2) |
| Other Sleep Disorders | ·· | 3613 (99) | 3613 (5) | ·· | 2876 (98) | 2876 (5) | ·· | 6489 (99) | 6489 (5) |
| Hypersomnia | ·· | 28 (1) | 28 (0) | ·· | 48 (2) | 48 (0) | ·· | 76 (1) | 76 (0) |
| Circadian Rhythm Sleep Disorders | ·· | 20 (1) | 20 (0) | ·· | 13 (0) | 13 (0) | ·· | 33 (1) | 33 (0) |
| Parasomnias | ·· | 168 (5) | 168 (0) | ·· | 77 (3) | 77 (0) | ·· | 245 (4) | 245 (0) |
| Non-Specific Sleep Disorders | ·· | 1686 (46) | 1686 (2) | ·· | 1003 (34) | 1003 (2) | ·· | 2689 (41) | 2689 (2) |
| Early Chronotype | 47971 (63) | 2172 (59) | 50143 (63) | 35981 (62) | 1796 (61) | 37777 (62) | 83952 (63) | 3968 (60) | 87920 (63) |
| Adequate Duration (7 to <9 hr/d) | 53284 (71) | 2138 (58) | 55422 (70) | 40824 (70) | 1713 (59) | 42537 (70) | 94108 (70) | 3851 (58) | 97959 (70) |
| No Usual Insomnia | 53176 (70) | 1902 (52) | 55078 (70) | 45630 (79) | 1873 (64) | 47503 (78) | 98806 (74) | 3775 (57) | 102581 (73) |
| No Snoring | 54559 (72) | 2338 (64) | 56897 (72) | 30380 (52) | 1178 (40) | 31558 (52) | 84939 (64) | 3516 (53) | 88455 (63) |
| No Frequent Daytime Sleepiness | 74051 (98) | 3500 (96) | 77551 (98) | 56669 (98) | 2737 (94) | 59406 (97) | 130720 (98) | 6237 (95) | 136957 (98) |
| Follow-up Time (yr) | 11·9 (1·1) | 11·8 (1·1) | 11·8 (1·1) | 11·8 (1·3) | 11·7 (1·3) | 11·8 (1·3) | 11·8 (1·2) | 11·7 (1·2) | 11·8 (1·2) |
| Incident CVD | 10277 (14) | 792 (22) | 11069 (14) | 11445 (20) | 966 (33) | 12411 (20) | 21722 (16) | 1758 (27) | 23480 (17) |
| All-Cause Death | 2479 (3) | 170 (5) | 2649 (3) | 3108 (5) | 239 (8) | 3347 (5) | 5587 (4) | 409 (6) | 5996 (4) |
| Age of Enrollment | 56·1 (8·0) | 56·1 (7·8) | 56·1 (8·0) | 56·2 (8·2) | 56·5 (8·1) | 56·2 (8·2) | 56·1 (8·1) | 56·3 (7·9) | 56·1 (8·1) |
| Sex | | | | | | | | | |
| Female | ·· | ·· | ·· | ·· | ·· | ·· | 75564 (57) | 3662 (56) | 79226 (57) |
| Male | ·· | ·· | ·· | ·· | ·· | ·· | 58030 (43) | 2925 (44) | 60955 (43) |
| Socioeconomic Status | | | | | | | | | |
| Highest Deprivation | 15835 (21) | 974 (27) | 16809 (21) | 12348 (21) | 797 (27) | 13145 (22) | 28183 (21) | 1771 (27) | 29954 (21) |
| 2nd Deprivation | 19552 (26) | 948 (26) | 20500 (26) | 14442 (25) | 734 (25) | 15176 (25) | 33994 (25) | 1682 (26) | 35676 (25) |
| 3rd Deprivation | 20003 (26) | 854 (23) | 20857 (26) | 15287 (26) | 698 (24) | 15985 (26) | 35290 (26) | 1552 (24) | 36842 (26) |
| Least Deprivation | 20174 (27) | 886 (24) | 21060 (27) | 15953 (27) | 696 (24) | 16649 (27) | 36127 (27) | 1582 (24) | 37709 (27) |
| Mental Health Issues | 30096 (40) | 2324 (63) | 32420 (41) | 14362 (25) | 1275 (44) | 15637 (26) | 44458 (33) | 3599 (55) | 48057 (34) |
| Perceived Health | | | | | | | | | |
| Excellent | 14944 (20) | 357 (10) | 15301 (19) | 10813 (19) | 276 (9) | 11089 (18) | 25757 (19) | 633 (10) | 26390 (19) |
| Good | 46721 (62) | 2061 (56) | 48782 (62) | 34801 (60) | 1470 (50) | 36271 (60) | 81522 (61) | 3531 (54) | 85053 (61) |

| | | | | | | | | | |
|---|---|---|---|---|---|---|---|---|---|
| Fair | 12103 (16) | 987 (27) | 13090 (17) | 10915 (19) | 917 (31) | 11832 (19) | 23018 (17) | 1904 (29) | 24922 (18) |
| Poor | 1796 (2) | 257 (7) | 2053 (3) | 1501 (3) | 262 (9) | 1763 (3) | 3297 (2) | 519 (8) | 3816 (3) |
| Body Mass Index (kg/m2) | | | | | | | | | |
| <30 | 59344 (79) | 2540 (69) | 61884 (78) | 44701 (77) | 1770 (61) | 46471 (76) | 104045 (78) | 4310 (65) | 108355 (77) |
| 30 to <40 | 14802 (20) | 914 (25) | 15716 (20) | 12818 (22) | 1009 (34) | 13827 (23) | 27620 (21) | 1923 (29) | 29543 (21) |
| >= 40 | 1418 (2) | 208 (6) | 1626 (2) | 511 (1) | 146 (5) | 657 (1) | 1929 (1) | 354 (5) | 2283 (2) |
| Economic Activity and Shift Work | | | | | | | | | |
| Retired/not in the workforce | 30796 (41) | 1696 (46) | 32492 (41) | 18918 (33) | 1135 (39) | 20053 (33) | 49714 (37) | 2831 (43) | 52545 (37) |
| Employed not in shift work | 38515 (51) | 1637 (45) | 40152 (51) | 31884 (55) | 1428 (49) | 33312 (55) | 70399 (53) | 3065 (47) | 73464 (52) |
| Employed in night shift work | 2648 (4) | 135 (4) | 2783 (4) | 4111 (7) | 194 (7) | 4305 (7) | 6759 (5) | 329 (5) | 7088 (5) |
| Employed in day shift work | 3605 (5) | 194 (5) | 3799 (5) | 3117 (5) | 168 (6) | 3285 (5) | 6722 (5) | 362 (5) | 7084 (5) |
| Cigarette Smoking | | | | | | | | | |
| Never | 46024 (61) | 2097 (57) | 48121 (61) | 30219 (52) | 1310 (45) | 31529 (52) | 76243 (57) | 3407 (52) | 79650 (57) |
| Cessation | 23504 (31) | 1166 (32) | 24670 (31) | 21153 (36) | 1234 (42) | 22387 (37) | 44657 (33) | 2400 (36) | 47057 (34) |
| Current | 6036 (8) | 399 (11) | 6435 (8) | 6658 (11) | 381 (13) | 7039 (12) | 12694 (10) | 780 (12) | 13474 (10) |
| Alcohol Drinking | | | | | | | | | |
| Never | 3591 (5) | 207 (6) | 3798 (5) | 1257 (2) | 68 (2) | 1325 (2) | 4848 (4) | 275 (4) | 5123 (4) |
| Quit | 2310 (3) | 179 (5) | 2489 (3) | 1620 (3) | 126 (4) | 1746 (3) | 3930 (3) | 305 (5) | 4235 (3) |
| < 14 UK Units/wk | 50732 (67) | 2433 (66) | 53165 (67) | 23997 (41) | 1211 (41) | 25208 (41) | 74729 (56) | 3644 (55) | 78373 (56) |
| 14 to <28 UK Units/wk | 13876 (18) | 571 (16) | 14447 (18) | 16026 (28) | 753 (26) | 16779 (28) | 29902 (22) | 1324 (20) | 31226 (22) |
| >= 28 UK Units/wk | 5055 (7) | 272 (7) | 5327 (7) | 15130 (26) | 767 (26) | 15897 (26) | 20185 (15) | 1039 (16) | 21224 (15) |
| Dietary Quality | | | | | | | | | |
| Poor | 2444 (3) | 151 (4) | 2595 (3) | 6083 (10) | 335 (11) | 6418 (11) | 8527 (6) | 486 (7) | 9013 (6) |
| Intermediate | 43400 (57) | 2145 (59) | 45545 (57) | 38021 (66) | 1919 (66) | 39940 (66) | 81421 (61) | 4064 (62) | 85485 (61) |
| Healthy | 29720 (39) | 1366 (37) | 31086 (39) | 13926 (24) | 671 (23) | 14597 (24) | 43646 (33) | 2037 (31) | 45683 (33) |
| Discretionary Screen Time (hr/d) | 3·6 (1·9) | 3·8 (2·1) | 3·6 (1·9) | 4·0 (2·1) | 4·4 (2·4) | 4·0 (2·1) | 3·8 (2·0) | 4·1 (2·3) | 3·8 (2·0) |
| Physical Activity | | | | | | | | | |
| No MVPA | 12504 (17) | 727 (20) | 13231 (17) | 8188 (14) | 530 (18) | 8718 (14) | 20692 (15) | 1257 (19) | 21949 (16) |
| < 10 MET hrs/wk | 7826 (10) | 403 (11) | 8229 (10) | 5900 (10) | 354 (12) | 6254 (10) | 13726 (10) | 757 (11) | 14483 (10) |
| 10 to <20 MET hrs/wk | 11874 (16) | 583 (16) | 12457 (16) | 8411 (14) | 417 (14) | 8828 (14) | 20285 (15) | 1000 (15) | 21285 (15) |
| >= 20 MET hrs/wk | 43360 (57) | 1949 (53) | 45309 (57) | 35531 (61) | 1624 (56) | 37155 (61) | 78891 (59) | 3573 (54) | 82464 (59) |

Data were shown in N (% vertically) or Mean (S.D.) for categorical or continuous variables, respectively.

**Table S10. Detailed Total, CVD-Free, with CVD Life Expectancy Estimation at Age 40**

| | Females | | | Males | | |
|---|---|---|---|---|---|---|
| Exposures | CVD-free | With CVD | Total | CVD-free | With CVD | Total |
| Total included participants | 33·05 [32·91-33·21] | 6·02 [5·86-6·16] | 39·07 [38·95-39·17] | 30·03 [29·88-30·20] | 8·26 [8·08-8·42] | 38·29 [38·16-38·41] |
| Primary Care Subsample | 33·30 [33·09-33·52] | 5·90 [5·67-6·11] | 39·20 [39·03-39·35] | 30·12 [29·90-30·37] | 8·16 [7·90-8·40] | 38·28 [38·09-38·46] |
| Sleep Characteristic | | | | | | |
| Usual Insomnia | 32·98 [32·67-33·28] | 6·31 [6·02-6·61] | 39·29 [39·09-39·46] | 30·09 [29·66-30·50] | 8·34 [7·93-8·75] | 38·44 [38·15-38·69] |
| No Usual | 33·28 [33·07-33·47] | 5·78 [5·58-5·99] | 39·06 [38·91-39·19] | 30·23 [30·01-30·45] | 7·97 [7·74-8·19] | 38·20 [38·04-38·36] |
| Inadequate Sleep Duration | 32·63 [32·32-32·93] | 6·39 [6·10-6·72] | 39·03 [38·82-39·22] | 29·86 [29·50-30·21] | 8·18 [7·82-8·52] | 38·05 [37·78-38·28] |
| Adequate | 33·26 [33·05-33·45] | 5·76 [5·56-5·97] | 39·02 [38·87-39·16] | 30·18 [29·94-30·42] | 7·98 [7·74-8·21] | 38·16 [37·98-38·33] |
| Snoring | 33·07 [32·76-33·37] | 6·19 [5·88-6·50] | 39·26 [39·05-39·44] | 30·06 [29·80-30·32] | 8·42 [8·15-8·69] | 38·48 [38·30-38·65] |
| No | 33·26 [33·06-33·45] | 5·78 [5·59-5·98] | 39·05 [38·90-39·18] | 30·41 [30·16-30·66] | 7·92 [7·67-8·17] | 38·34 [38·16-38·50] |
| Evening Chronotype | 33·21 [32·93-33·47] | 5·74 [5·47-6·01] | 38·95 [38·75-39·14] | 30·20 [29·90-30·51] | 7·84 [7·52-8·15] | 38·04 [37·81-38·26] |
| Morning | 33·20 [32·99-33·39] | 5·85 [5·64-6·06] | 39·05 [38·89-39·19] | 30·29 [30·05-30·52] | 7·99 [7·74-8·22] | 38·27 [38·10-38·44] |
| Frequent Daytime Sleepiness | 32·33 [31·00-33·37] | 6·86 [5·71-8·07] | 39·19 [38·36-39·72] | 29·85 [28·40-30·98] | 8·36 [7·00-9·66] | 38·20 [37·06-38·96] |
| Infrequent | 33·24 [33·06-33·41] | 5·79 [5·61-5·96] | 39·03 [38·91-39·15] | 30·26 [30·05-30·46] | 7·99 [7·76-8·19] | 38·24 [38·09-38·37] |
| Composite Sleep Score | | | | | | |
| ≤ 1 | 31·46 [30·36-32·48] | 7·72 [6·61-8·76] | 39·18 [38·28-39·72] | 27·96 [26·80-29·02] | 10·17 [8·95-11·34] | 38·13 [37·03-38·87] |
| 2 | 32·73 [32·27-33·19] | 6·37 [5·88-6·79] | 39·10 [38·72-39·41] | 29·36 [28·86-29·86] | 9·06 [8·52-9·56] | 38·42 [38·04-38·74] |
| 3 | 32·78 [32·51-33·09] | 6·26 [5·96-6·54] | 39·05 [38·82-39·25] | 29·84 [29·57-30·15] | 8·41 [8·09-8·69] | 38·25 [38·01-38·46] |
| 4 | 33·32 [33·09-33·58] | 5·93 [5·67-6·15] | 39·25 [39·06-39·41] | 30·30 [30·06-30·57] | 8·10 [7·81-8·35] | 38·40 [38·19-38·59] |
| 5 | 33·36 [33·07-33·68] | 5·63 [5·29-5·93] | 38·99 [38·72-39·22] | 30·17 [29·81-30·56] | 8·04 [7·61-8·42] | 38·22 [37·87-38·50] |
| Poor (≤ 1) | 31·46 [30·36-32·48] | 7·72 [6·61-8·76] | 39·18 [38·28-39·72] | 27·96 [26·80-29·02] | 10·17 [8·95-11·34] | 38·13 [37·03-38·87] |
| Intermediate (2 To 3) | 32·77 [32·54-33·03] | 6·30 [6·04-6·52] | 39·07 [38·88-39·24] | 29·72 [29·48-29·97] | 8·54 [8·27-8·79] | 38·26 [38·06-38·45] |
| Healthy (≥ 4) | 33·26 [33·08-33·46] | 5·77 [5·57-5·95] | 39·03 [38·87-39·17] | 30·27 [30·07-30·49] | 8·02 [7·78-8·23] | 38·29 [38·11-38·45] |
| Diagnosed Sleep Disorder | | | | | | |
| Insomnia | 31·77 [28·23-33·78] | 6·23 [3·98-8·59] | 38·00 [34·36-39·30] | 26·32 [21·34-29·73] | 13·46 [8·54-17·64] | 39·78 [34·81-40·68] |
| Without This Condition | 33·36 [33·11-33·57] | 5·88 [5·67-6·09] | 39·24 [39·06-39·38] | 30·16 [29·92-30·39] | 8·07 [7·84-8·31] | 38·23 [38·03-38·41] |
| Sleep-Related Breathing Disorders | 26·01 [22·80-28·21] | 10·20 [7·48-12·54] | 36·22 [32·27-38·09] | 23·48 [21·54-25·10] | 12·77 [10·67-14·76] | 36·25 [34·14-37·61] |
| Without This Condition | 33·34 [33·09-33·56] | 5·90 [5·69-6·12] | 39·24 [39·07-39·38] | 30·21 [29·96-30·44] | 8·05 [7·81-8·29] | 38·26 [38·05-38·43] |
| Other Sleep Disorders | 31·91 [29·81-33·34] | 7·50 [5·83-9·14] | 39·41 [37·92-40·05] | 29·19 [27·11-30·77] | 8·59 [6·69-10·39] | 37·78 [35·78-38·87] |
| Without This Condition | 33·33 [33·09-33·55] | 5·88 [5·67-6·10] | 39·21 [39·04-39·36] | 30·18 [29·93-30·41] | 8·07 [7·84-8·31] | 38·25 [38·05-38·43] |
| With Any Sleep Disorders | 30·64 [29·46-31·74] | 7·65 [6·44-8·69] | 38·29 [37·08-39·06] | 26·46 [25·30-27·58] | 10·67 [9·27-11·96] | 37·12 [35·91-38·02] |
| Without Sleep Disorders | 33·37 [33·16-33·60] | 5·85 [5·61-6·06] | 39·22 [39·05-39·37] | 30·24 [30·01-30·50] | 8·05 [7·77-8·29] | 38·29 [38·09-38·47] |

Data were shown in life expectancy estimation [95% CI]. For all the estimation, we applied median or mean value for categorical or numerous covariates, respectively, and for estimations of each self-reported sleep characteristic or each diagnosed sleep disorder, the remaining characteristics or disorders were set to healthy.

**Table S11. Total Life Expectancy and Years of Life Lost at Age 40 among Participants with Different Sleep Exposures**

| | Females | | Males | |
|---|---|---|---|---|
| Exposures | Total Life Expectancy | Total Years of Life Lost | Total Life Expectancy | Total Years of Life Lost |
| Total included participants | 39·07 [38·95-39·17] | ·· | 38·29 [38·16-38·41] | ·· |
| Primary Care Subsample | 39·20 [39·03-39·35] | ·· | 38·28 [38·09-38·46] | ·· |
| Sleep Characteristic* | | | | |
| Usual Insomnia | 39·29 [39·09-39·46] | **-0·23 [-0·29--0·15]** | 38·44 [38·15-38·69] | **-0·24 [-0·35--0·11]** |
| No Usual Insomnia | 39·06 [38·91-39·19] | ·· | 38·20 [38·04-38·36] | ·· |
| Inadequate Sleep Duration | 39·03 [38·82-39·22] | 0·00 [-0·07-0·08] | 38·05 [37·78-38·28] | **0·11 [0·04-0·21]** |
| Adequate Duration | 39·02 [38·87-39·16] | ·· | 38·16 [37·98-38·33] | ·· |
| Snoring | 39·26 [39·05-39·44] | **-0·21 [-0·27-0·14]** | 38·48 [38·30-38·65] | **-0·15 [-0·17-0·12]** |
| No Snoring | 39·05 [38·90-39·18] | ·· | 38·34 [38·16-38·50] | ·· |
| Evening Chronotype | 38·95 [38·75-39·14] | **0·09 [0·04-0·15]** | 38·04 [37·81-38·26] | **0·23 [0·17-0·30]** |
| Morning Chronotype | 39·05 [38·89-39·19] | ·· | 38·27 [38·10-38·44] | ·· |
| Frequent Daytime Sleepiness | 39·19 [38·36-39·72] | 0·15 [-0·57-0·55] | 38·20 [37·06-38·96] | 0·04 [-0·59-1·02] |
| Infrequent Daytime Sleepiness | 39·03 [38·91-39·15] | ·· | 38·24 [38·09-38·37] | ·· |
| Composite Sleep Score† | | | | |
| ≤ 1 | 39·18 [38·28-39·72] | 0·19 [-0·56-0·51] | 38·13 [37·03-38·87] | 0·08 [-0·41-0·90] |
| 2 | 39·10 [38·72-39·41] | 0·11 [-0·23-0·04] | 38·42 [38·04-38·74] | **-0·20 [-0·31--0·07]** |
| 3 | 39·05 [38·82-39·25] | 0·06 [-0·13-0·01] | 38·25 [38·01-38·46] | -0·04 [-0·16-0·06] |
| 4 | 39·25 [39·06-39·41] | **-0·26 [-0·34--0·18]** | 38·40 [38·19-38·59] | **-0·18 [-0·32--0·08]** |
| 5 | 38·99 [38·72-39·22] | ·· | 38·22 [37·87-38·50] | ·· |
| Poor (≤ 1) | 39·18 [38·28-39·72] | -0·15 [-0·58-0·62] | 38·13 [37·03-38·87] | 0·16 [-0·43-1·12] |
| Intermediate (2 to 3) | 39·07 [38·88-39·24] | -0·04 [-0·08-0·01] | 38·26 [38·06-38·45] | 0·03 [-0·01-0·07] |
| Healthy (≥ 4) | 39·03 [38·87-39·17] | ·· | 38·29 [38·11-38·45] | ·· |
| Clinical Diagnosed Sleep Disorder* | | | | |
| Insomnia | 38·00 [34·36-39·30] | **1·23 [0·06-4·73]** | 39·78 [34·81-40·68] | -1·55 [-2·31-3·27] |
| Without This Condition | 39·24 [39·06-39·38] | ·· | 38·23 [38·03-38·41] | ·· |
| Sleep-Related Breathing Disorders | 36·22 [32·27-38·09] | **3·02 [1·29-6·81]** | 36·25 [34·14-37·61] | **2·01 [0·77-3·93]** |
| Without This Condition | 39·24 [39·07-39·38] | ·· | 38·26 [38·05-38·43] | ·· |
| Other Sleep Disorders | 39·41 [37·92-40·05] | 0·19 [-0·71-1·14] | 37·78 [35·78-38·87] | 0·47 [-0·47-2·28] |
| Without This Condition | 39·21 [39·04-39·36] | ·· | 38·25 [38·05-38·43] | ·· |
| With Any Sleep Disorders | 38·29 [37·08-39·06] | **0·93 [0·31-1·97]** | 37·12 [35·91-38·02] | **1·16 [0·43-2·21]** |
| Without Any Sleep Disorders | 39·22 [39·05-39·37] | ·· | 38·29 [38·09-38·47] | ·· |

Data were shown in estimation [95% CI], with values in bold denoting statistical significance. For all the estimation, we applied median or mean value for categorical or numerous covariates, respectively, and for estimations of each self-reported sleep characteristic or each diagnosed sleep disorder, the remaining characteristics or disorders were set to healthy. *Compared to those without poor sleep characteristics/diagnosed conditions.

**Table S12. Confusion Matrix of Self-Reported Insomnia/Snoring with Diagnosed Insomnia/Sleep-Related Breathing Disorders**

| | Diagnosed Insomnia | | | | Diagnosed Sleep-Related Breathing | | |
|---|---|---|---|---|---|---|---|
| Self-reported* | Yes | No | Total | Self-reported† | Yes | No | Total |
| Usual Insomnia | 748 | 36852 | 37600 | Snoring | 1609 | 50117 | 51726 |
| No Usual Insomnia | 455 | 102126 | 102581 | No Snoring | 832 | 87623 | 88455 |
| Total | 1203 | 138978 | 140181 | Total | 2441 | 138042 | 140181 |

*Sensitivity: 0·62 [0·59, 0·65]; Specificity: 0·73 [0·73, 0·74]; Positive Predictive Value, 0·02 [0·02, 0·02]; Negative Predictive Value: 1·00 [1·00, 1·00].

†Sensitivity: 0·66 [0·64, 0·68]; Specificity: 0·64 [0·63, 0·64]; Positive Predictive Value, 0·03 [0·03, 0·03]; Negative Predictive Value: 0·99 [0·99, 0·99].

#### Supplemental Figures

**Figure S1. Primary Care Clinical Events Data Flowchart**

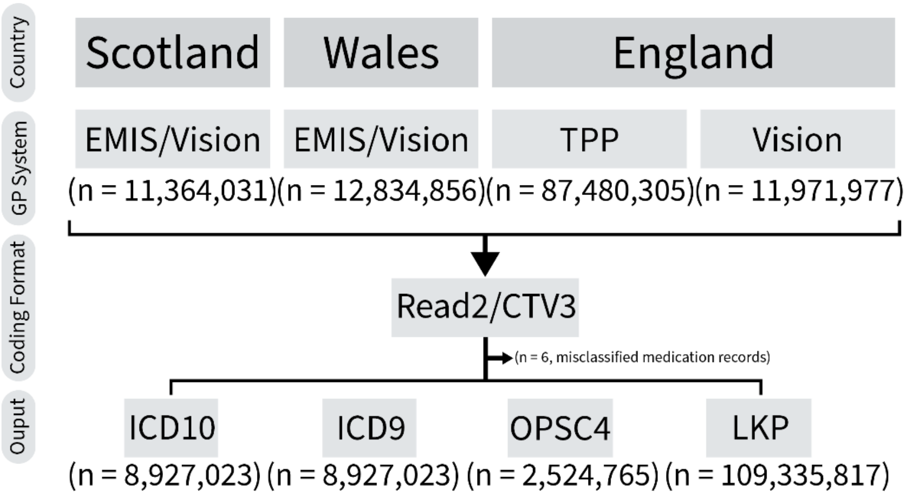

Abbreviations: CTV, Clinical Terms Version; ICD, International Classification of Diseases; LKP, directly LooK-Up; OPCS, Office of Population Censuses and Surveys Classification of Surgical Operations and Procedures.

**Figure S2. Primary Care Prescription Data Flowchart**

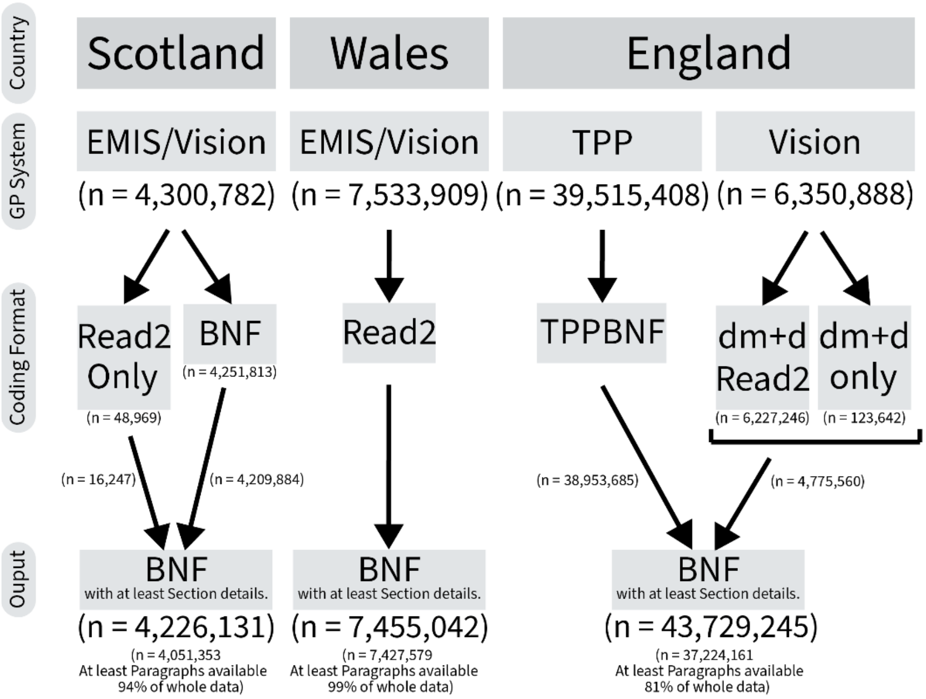

Abbreviations: BNF, British National Formulary; dm+d, Dictionary of Medicines and Devices; EMIS, the Egton Medical Information Systems health; Read2, Read codes version 2; TPP, The Phoenix Partnership; Vision, the Vision health.

**Figure S3. *a priori* Defined Directed Acyclic Graphs**

(A)

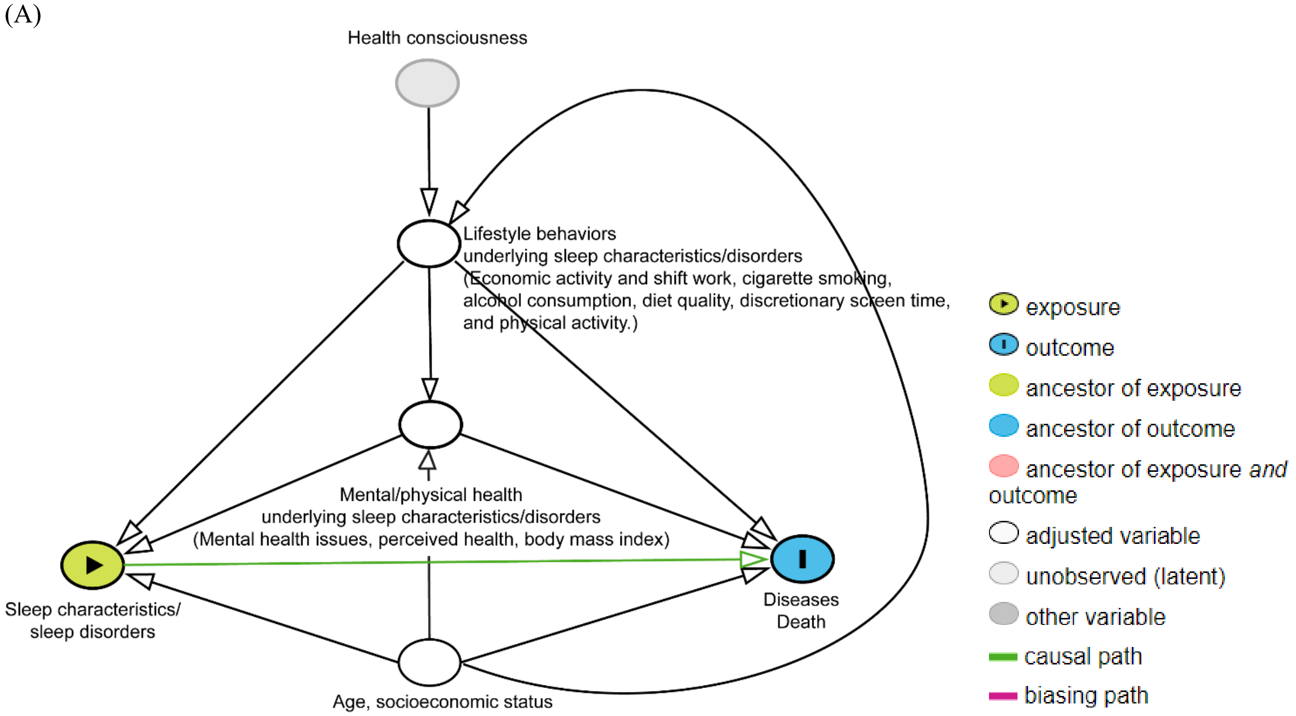

(B)

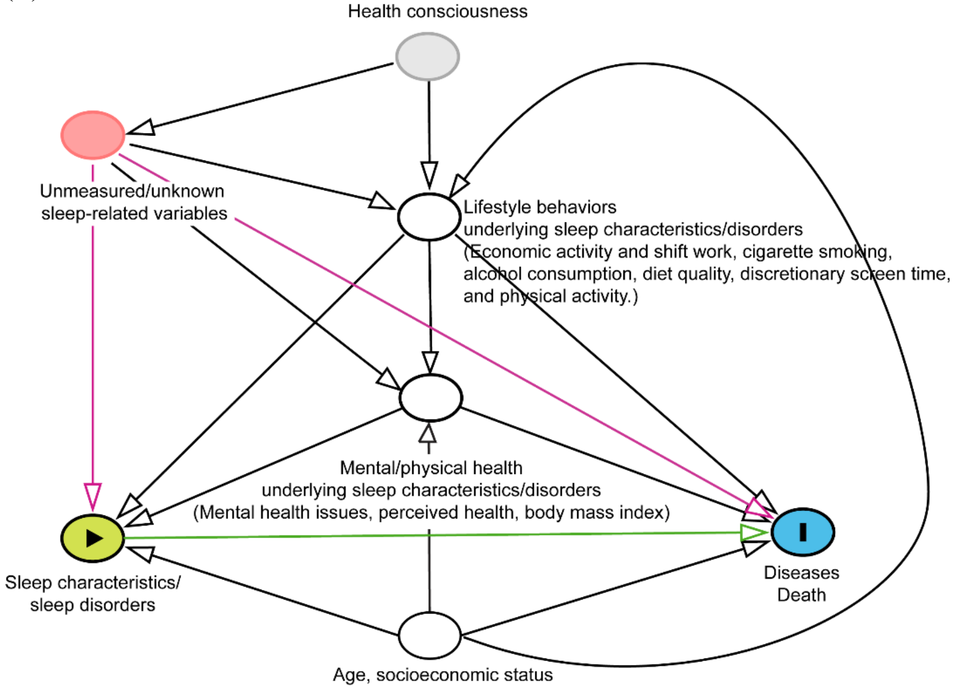

(A) By controlling lifestyle behaviors and mental/physical health underlying sleep, we could block the backdoor path introduced by the unmeasurable confounder “health consciousness”. Age served as a time-varying variable in the life expectancy estimation. We also controlled socioeconomic status with rationales explained in B. (B) The potential unmeasured or unknown sleep-related variables remains as a theoretical limitation of observational studies. Although by conditioning on age and socioeconomic status, we could avoid the potential backdoor path introduced by the colliders of lifestyle behaviors and mental/physical health underlying sleep, we still could not fully rule out the potential direct biasing path via the unmeasured/unknown variables.

The directed acyclic graphs above were generated via an open-source application (Johannes Textor, Benito van der Zander, Mark K. Gilthorpe, Maciej Liskiewicz, George T.H. Ellison. Robust causal inference using directed acyclic graphs: the R package 'dagitty'. *International Journal of Epidemiology* 45(6):1887-1894, 2016. https://doi.org/10.1093/ije/dyw341), and further modified by Adobe Illustrator (Adobe Inc., 2019., available at: https://adobe.com/products/illustrator.)

**Figure S4. Total and Cardiovascular Disease-Free Life Expectancy at Age 40 among Participants with Self-Reported Sleep Characteristics**

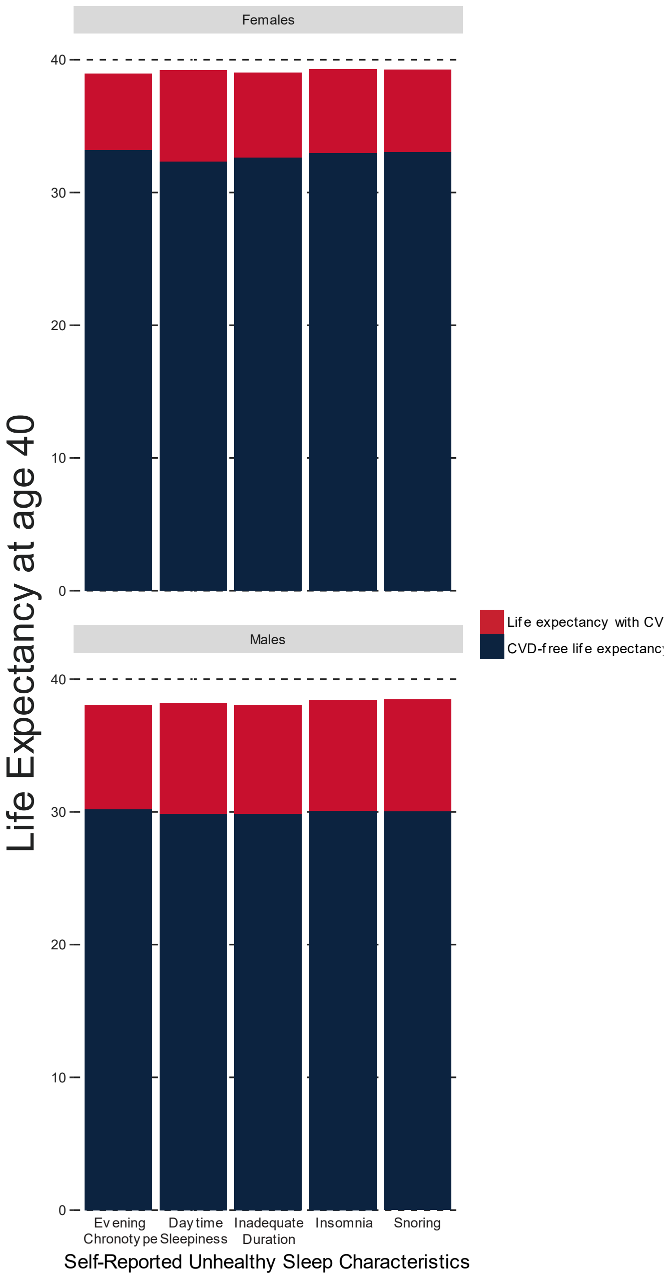

**Figure S5. Risks for Cardiovascular Diseases between Participants with Different Self-Reported Sleep Characteristic**

(A) Females with different composite sleep score

| Variables | Hazard Ratio (HR) HR [95% CI] |
|---|---|
| **Sleep** | |
| Poor | 1.13 [1.05−1.22] |
| Intermediate | 1.05 [1.02−1.08] |
| Healthy | 1 |
| **Age of Enrolment** | |
| Each year older | 1.07 [1.06−1.07] |
| **Socioeconomic Status** | |
| Highest Deprivation | 1.09 [1.05−1.14] |
| 2nd Deprivation | 1.04 [1−1.08] |
| 3rd Deprivation | 1.04 [1.01−1.08] |
| Least Deprivation | 1 |
| **Mental Health Issues** | |
| Yes | 1.11 [1.08−1.13] |
| No | 1 |
| **Perceived Health** | |
| Poor | 2.73 [2.53−2.94] |
| Fair | 2.04 [1.95−2.13] |
| Good | 1.43 [1.38−1.49] |
| Excellent | 1 |
| **BodyMass Index (kg/m2)** | |
| >= 40 | 1.48 [1.38−1.59] |
| 30 to <40 | 1.13 [1.1−1.17] |
| <30 | 1 |
| **Economic Activityand Shift Work** | |
| Employed in night shift work | 1.04 [0.96−1.13] |
| Employed in day shift work | 1.04 [0.97−1.11] |
| Employed not in shift work | 0.94 [0.91−0.97] |
| Retired/not in the workforce | 1 |
| **Cigarette Smoking** | |
| Current | 1.2 [1.15−1.26] |
| Cessation | 1.11 [1.08−1.14] |
| Never | 1 |
| **Alcohol Drinking** | |
| Quit | 1.03 [0.95−1.11] |
| Occational | 0.9 [0.85−0.95] |
| < 14 UK Units/wk | 0.84 [0.79−0.89] |
| 14 to <28 UK Units/wk | 0.81 [0.77−0.87] |
| >= 28 UK Units/wk | 0.86 [0.8−0.92] |
| Never | 1 |
| **DietaryQuality** | |
| Poor | 1.09 [1.02−1.17] |
| Intermediate | 1.03 [1−1.05] |
| Healthy | 1 |
| **DiscretionaryScreen Time (hr/d)** | |
| Each hour more | 1.03 [1.02−1.04] |
| **Physical Activity** | |
| No MVPA | 0.97 [0.94−1.01] |
| < 10 MET hrs/wk | 0.92 [0.88−0.96] |
| 10 to <20 MET hrs/wk | 0.94 [0.91−0.97] |
| >= 20 MET hrs/wk | 1 |

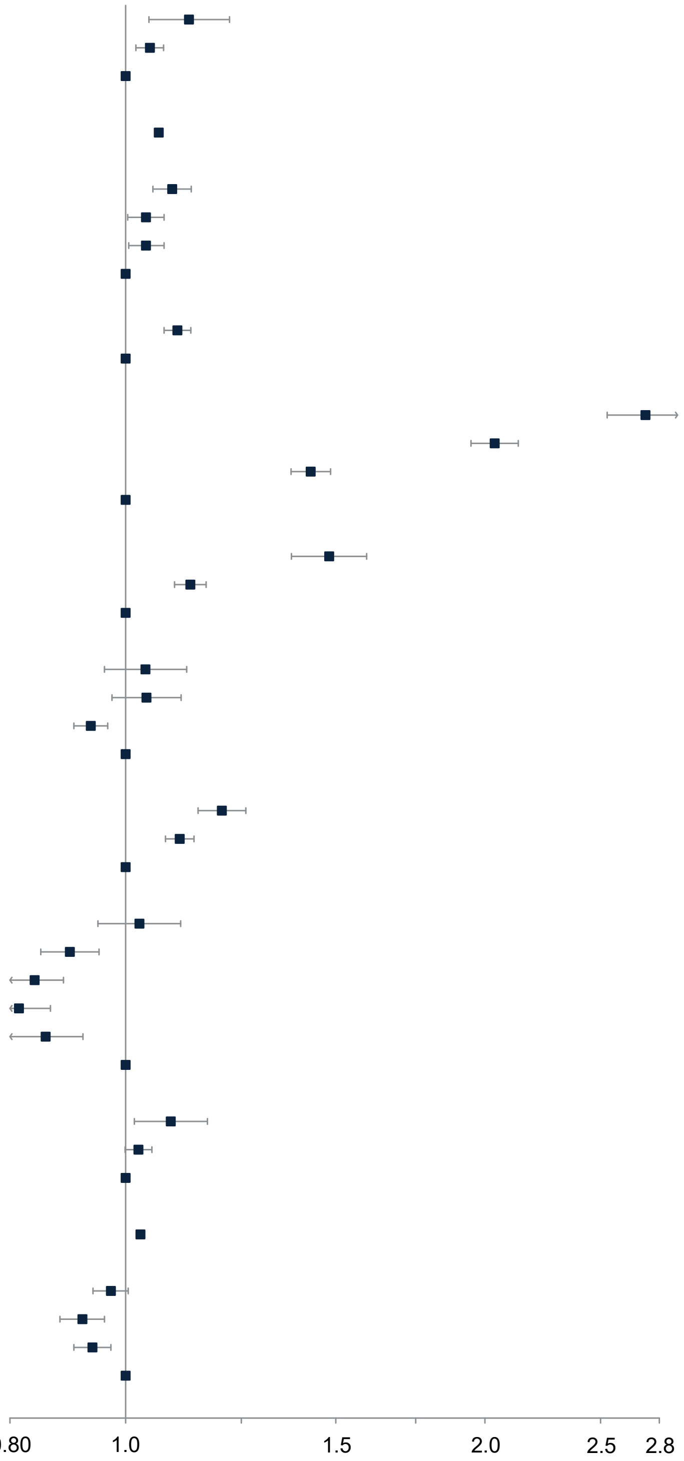

(B) Males with different composite sleep score

| Variables | Hazard Ratio (HR) HR [95% CI] |
|---|---|
| **Sleep** | |
| Poor | 1.17 [1.09−1.26] |
| Intermediate | 1.05 [1.02−1.07] |
| Healthy | 1 |
| **Age of Enrolment** | |
| Each year older | 1.07 [1.07−1.07] |
| **Socioeconomic Status** | |
| Highest Deprivation | 1.08 [1.04−1.12] |
| 2nd Deprivation | 1.04 [1.01−1.08] |
| 3rd Deprivation | 1.02 [0.99−1.05] |
| Least Deprivation | 1 |
| **Mental Health Issues** | |
| Yes | 1.04 [1.01−1.07] |
| No | 1 |
| **Perceived Health** | |
| Poor | 2.1 [1.96−2.26] |
| Fair | 1.69 [1.62−1.76] |
| Good | 1.29 [1.24−1.33] |
| Excellent | 1 |
| **Body Mass Index (kg/m2)** | |
| >= 40 | 1.61 [1.46−1.76] |
| 30 to <40 | 1.19 [1.16−1.23] |
| <30 | 1 |
| **Economic Activity and Shift Work** | |
| Employed in night shift work | 1.01 [0.96−1.07] |
| Employed in day shift work | 0.97 [0.91−1.03] |
| Employed not in shift work | 0.93 [0.9−0.96] |
| Retired/not in the workforce | 1 |
| **Cigarette Smoking** | |
| Current | 1.23 [1.18−1.28] |
| Cessation | 1.06 [1.04−1.09] |
| Never | 1 |
| **Alcohol Drinking** | |
| Quit | 1.12 [1.01−1.24] |
| Occasional | 1.05 [0.97−1.15] |
| < 14 UK Units/wk | 0.97 [0.89−1.05] |
| 14 to <28 UK Units/wk | 0.95 [0.87−1.04] |
| >= 28 UK Units/wk | 0.99 [0.91−1.08] |
| Never | 1 |
| **Dietary Quality** | |
| Poor | 0.98 [0.94−1.03] |
| Intermediate | 0.99 [0.96−1.02] |
| Healthy | 1 |
| **Discretionary Screen Time (hr/d)** | |
| Each hour more | 1.02 [1.01−1.03] |
| **Physical Activity** | |
| No MVPA | 0.94 [0.9−0.97] |
| < 10 MET hrs/wk | 0.95 [0.91−0.99] |
| 10 to <20 MET hrs/wk | 0.94 [0.91−0.97] |
| >= 20 MET hrs/wk | 1 |

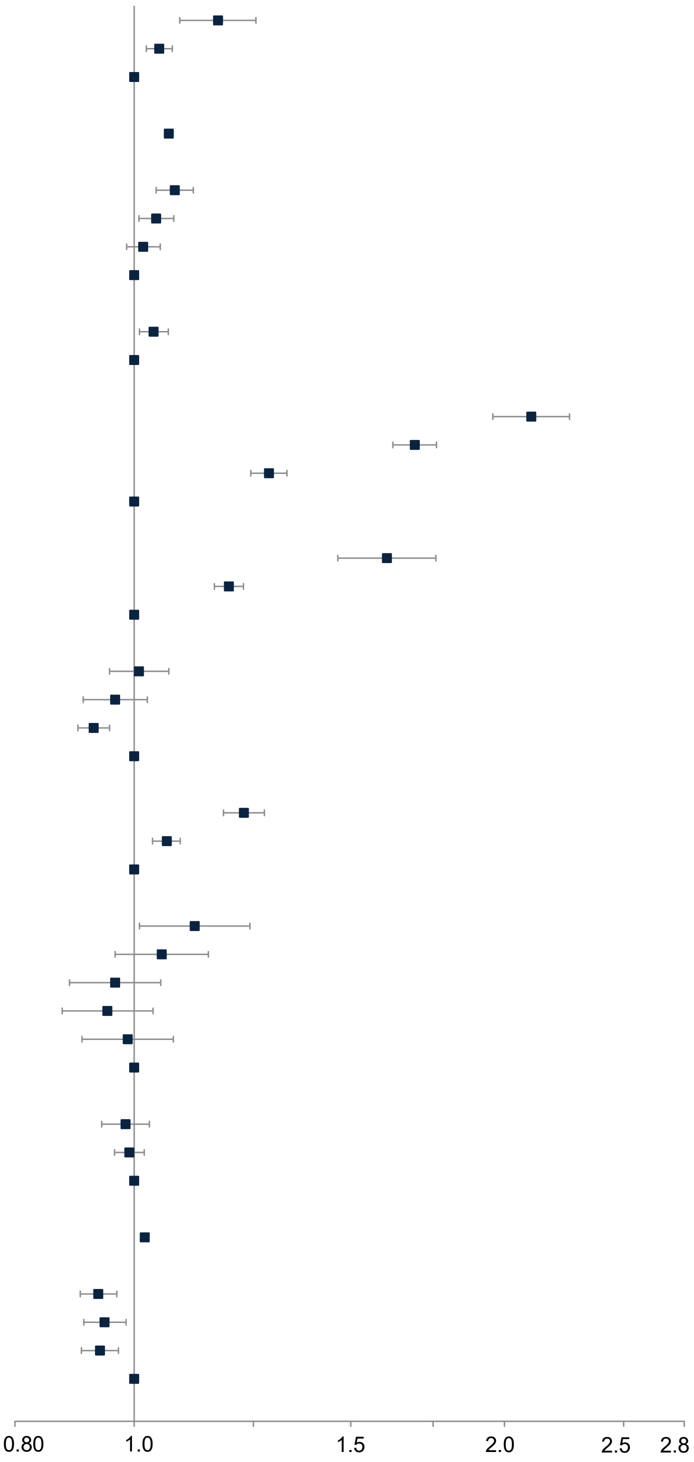

(C) Females with different sleep characteristics

| Variables | Hazard Ratio (HR) HR [95% CI] |
|---|---|
| **Chronotype** | |
| Evening | 0.99 [0.96−1.01] |
| Early | 1 |
| **Sleep Duration** | |
| Inadequate | 1.06 [1.03−1.09] |
| Adequate (7 to <9 hr/d) | 1 |
| **Insomnia** | |
| Usual | 1.03 [1−1.06] |
| Not usual | 1 |
| **Snoring** | |
| Yes | 1.03 [1−1.06] |
| No | 1 |
| **Daytime Sleepiness** | |
| Frequent | 1.11 [1.03−1.2] |
| Not frequent | 1 |
| **Age of Enrolment** | |
| Each year older | 1.07 [1.06−1.07] |
| **Socioeconomic Status** | |
| Highest Deprivation | 1.09 [1.05−1.13] |
| 2nd Deprivation | 1.04 [1−1.08] |
| 3rd Deprivation | 1.04 [1.01−1.08] |
| Least Deprivation | 1 |
| **Mental Health Issues** | |
| Yes | 1.1 [1.08−1.13] |
| No | 1 |
| **Perceived Health** | |
| Poor | 2.7 [2.51−2.91] |
| Fair | 2.03 [1.94−2.13] |
| Good | 1.43 [1.38−1.48] |
| Excellent | 1 |
| **Body Mass Index (kg/m2)** | |
| >= 40 | 1.48 [1.37−1.59] |
| 30 to <40 | 1.13 [1.1−1.17] |
| <30 | 1 |
| **Economic Activity and Shift Work** | |
| Employed in night shift work | 1.04 [0.96−1.13] |
| Employed in day shift work | 1.04 [0.97−1.11] |
| Employed not in shift work | 0.94 [0.91−0.97] |
| Retired/not in the workforce | 1 |
| **Cigarette Smoking** | |
| Current | 1.21 [1.16−1.27] |
| Cessation | 1.11 [1.08−1.14] |
| Never | 1 |
| **Alcohol Drinking** | |
| Quit | 1.03 [0.95−1.11] |
| Occasional | 0.9 [0.85−0.95] |
| < 14 UK Units/wk | 0.84 [0.8−0.89] |
| 14 to <28 UK Units/wk | 0.82 [0.77−0.87] |
| >= 28 UK Units/wk | 0.86 [0.8−0.93] |
| Never | 1 |
| **Dietary Quality** | |
| Poor | 1.09 [1.02−1.17] |
| Intermediate | 1.03 [1−1.05] |
| Healthy | 1 |
| **Discretionary Screen Time (hr/d)** | |
| Each hour more | 1.03 [1.02−1.04] |
| **Physical Activity** | |
| No MVPA | 0.97 [0.94−1.01] |
| < 10 MET hrs/wk | 0.92 [0.88−0.96] |
| 10 to <20 MET hrs/wk | 0.94 [0.91−0.97] |
| >= 20 MET hrs/wk | 1 |

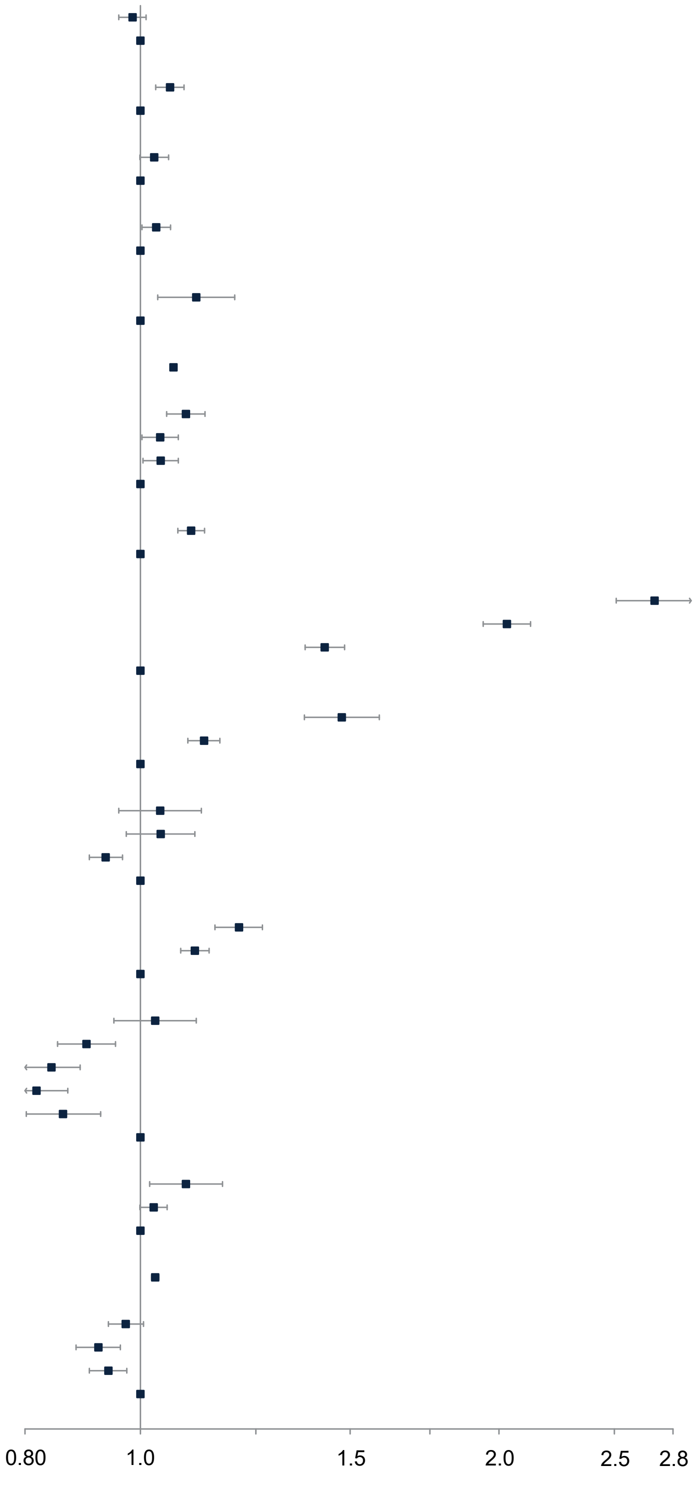

(D) Males with different sleep characteristics

| Variables | Hazard Ratio (HR) HR [95% CI] |
|---|---|
| **Chronotype** | |
| Evening | 1.01 [0.98−1.03] |
| Early | 1 |
| **Sleep Duration** | |
| Inadequate | 1.05 [1.02−1.07] |
| Adequate (7 to <9 hr/d) | 1 |
| **Insomnia** | |
| Usual | 1.03 [1−1.06] |
| Not usual | 1 |
| **Snoring** | |
| Yes | 1.03 [1.01−1.06] |
| No | 1 |
| **Daytime Sleepiness** | |
| Frequent | 1.06 [0.99−1.13] |
| Not frequent | 1 |
| **Age of Enrolment** | |
| Each year older | 1.07 [1.07−1.07] |
| **Socioeconomic Status** | |
| Highest Deprivation | 1.08 [1.03−1.12] |
| 2nd Deprivation | 1.04 [1−1.08] |
| 3rd Deprivation | 1.02 [0.99−1.05] |
| Least Deprivation | 1 |
| **Mental Health Issues** | |
| Yes | 1.04 [1.01−1.07] |
| No | 1 |
| **Perceived Health** | |
| Poor | 2.1 [1.98−2.26] |
| Fair | 1.69 [1.62−1.76] |
| Good | 1.29 [1.25−1.33] |
| Excellent | 1 |
| **Body Mass Index (kg/m2)** | |
| >= 40 | 1.6 [1.45−1.76] |
| 30 to <40 | 1.19 [1.15−1.23] |
| <30 | 1 |
| **Economic Activity and Shift Work** | |
| Employed in night shift work | 1.01 [0.97−1.07] |
| Employed in day shift work | 0.96 [0.93−1.02] |
| Employed not in shift work | 0.93 [0.92−0.95] |
| Retired/not in the workforce | 1 |
| **Cigarette Smoking** | |
| Current | 1.23 [1.15−1.28] |
| Cessation | 1.06 [1.03−1.09] |
| Never | 1 |
| **Alcohol Drinking** | |
| Quit | 1.12 [1.01−1.24] |
| Occasional | 1.05 [0.96−1.15] |
| < 14 UK Units/wk | 0.97 [0.88−1.05] |
| 14 to <28 UK Units/wk | 0.95 [0.87−1.04] |
| >= 28 UK Units/wk | 0.99 [0.9−1.08] |
| Never | 1 |
| **Dietary Quality** | |
| Poor | 0.99 [0.93−1.03] |
| Intermediate | 0.99 [0.96−1.02] |
| Healthy | 1 |
| **Discretionary Screen Time (hr/d)** | |
| Each hour more | 1.02 [1.02−1.03] |
| **Physical Activity** | |
| No MVPA | 0.94 [0.9−0.97] |
| < 10 MET hrs/wk | 0.95 [0.91−0.99] |
| 10 to <20 MET hrs/wk | 0.94 [0.91−0.97] |
| >= 20 MET hrs/wk | 1 |

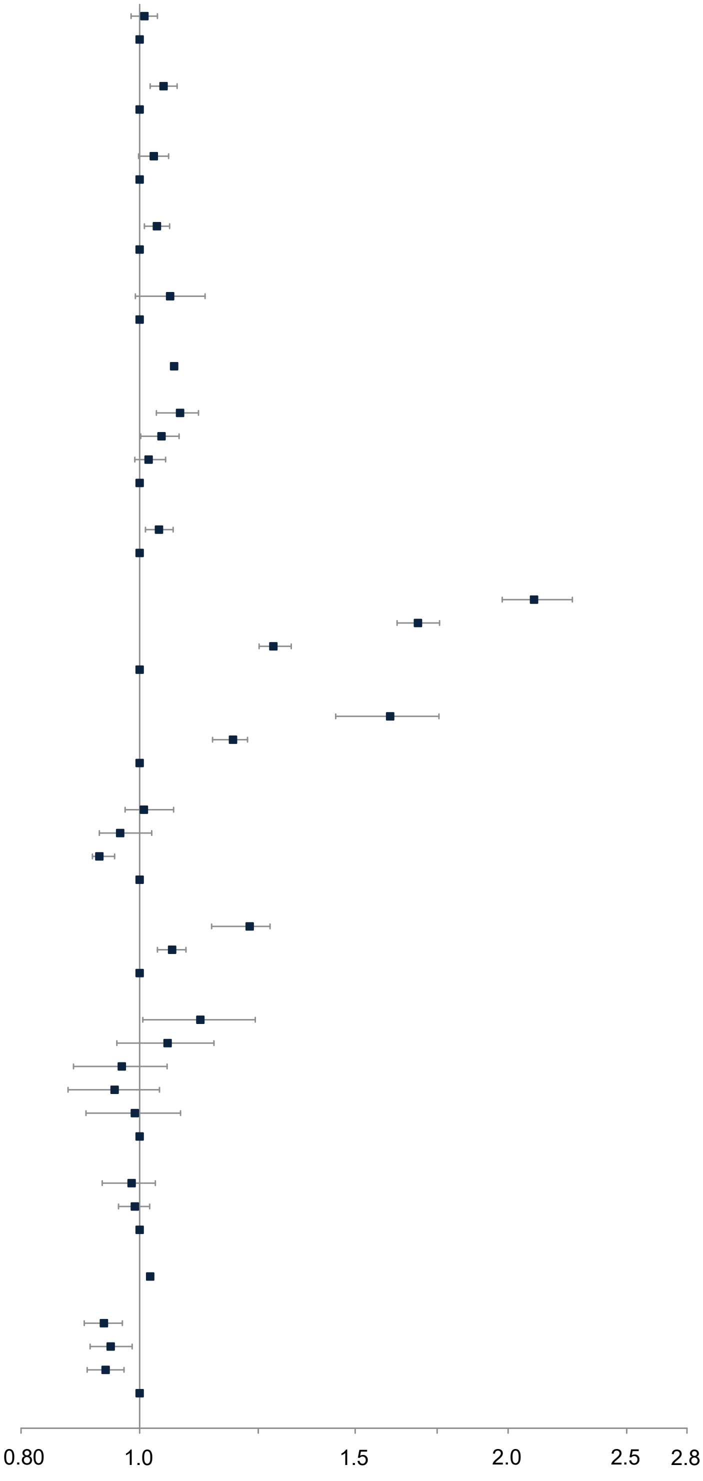

**Figure S6. Risks for Cardiovascular Diseases between Participants with Different Diagnosed Sleep Disorders**

(A) Females with any diagnosed sleep disorders

| Variables | Hazard Ratio (HR) HR [95% CI] |
|---|---|
| **Any Diagnosed Sleep Disorder** | |
| Yes | 1.51 [1.41−1.63] |
| No | 1 |
| **Age of Enrolment** | |
| Each year older | 1.07 [1.06−1.07] |
| **Socioeconomic Status** | |
| Highest Deprivation | 1.08 [1.02−1.14] |
| 2nd Deprivation | 1.05 [1−1.11] |
| 3rd Deprivation | 1.05 [1−1.11] |
| Least Deprivation | 1 |
| **Mental Health Issues** | |
| Yes | 1.11 [1.07−1.16] |
| No | 1 |
| **Perceived Health** | |
| Poor | 2.84 [2.55−3.17] |
| Fair | 2.05 [1.92−2.2] |
| Good | 1.46 [1.38−1.54] |
| Excellent | 1 |
| **Body Mass Index (kg/m2)** | |
| >= 40 | 1.53 [1.38−1.69] |
| 30 to <40 | 1.14 [1.09−1.2] |
| <30 | 1 |
| **Economic Activity and Shift Work** | |
| Employed in night shift work | 1 [0.89−1.13] |
| Employed in day shift work | 1 [0.91−1.11] |
| Employed not in shift work | 0.94 [0.89−0.99] |
| Retired/not in the workforce | 1 |
| **Cigarette Smoking** | |
| Current | 1.24 [1.16−1.33] |
| Cessation | 1.12 [1.08−1.17] |
| Never | 1 |
| **Alcohol Drinking** | |
| Quit | 1.04 [0.92−1.17] |
| Occasional | 0.91 [0.83−0.98] |
| < 14 UK Units/wk | 0.87 [0.8−0.94] |
| 14 to <28 UK Units/wk | 0.84 [0.77−0.92] |
| >= 28 UK Units/wk | 0.87 [0.78−0.97] |
| Never | 1 |
| **Dietary Quality** | |
| Poor | 1.07 [0.97−1.19] |
| Intermediate | 1.02 [0.98−1.06] |
| Healthy | 1 |
| **Discretionary Screen Time (hr/d)** | |
| Each hour more | 1.02 [1.01−1.03] |
| **Physical Activity** | |
| No MVPA | 0.96 [0.91−1.01] |
| < 10 MET hrs/wk | 0.93 [0.87−0.99] |
| 10 to <20 MET hrs/wk | 0.93 [0.88−0.98] |
| >= 20 MET hrs/wk | 1 |

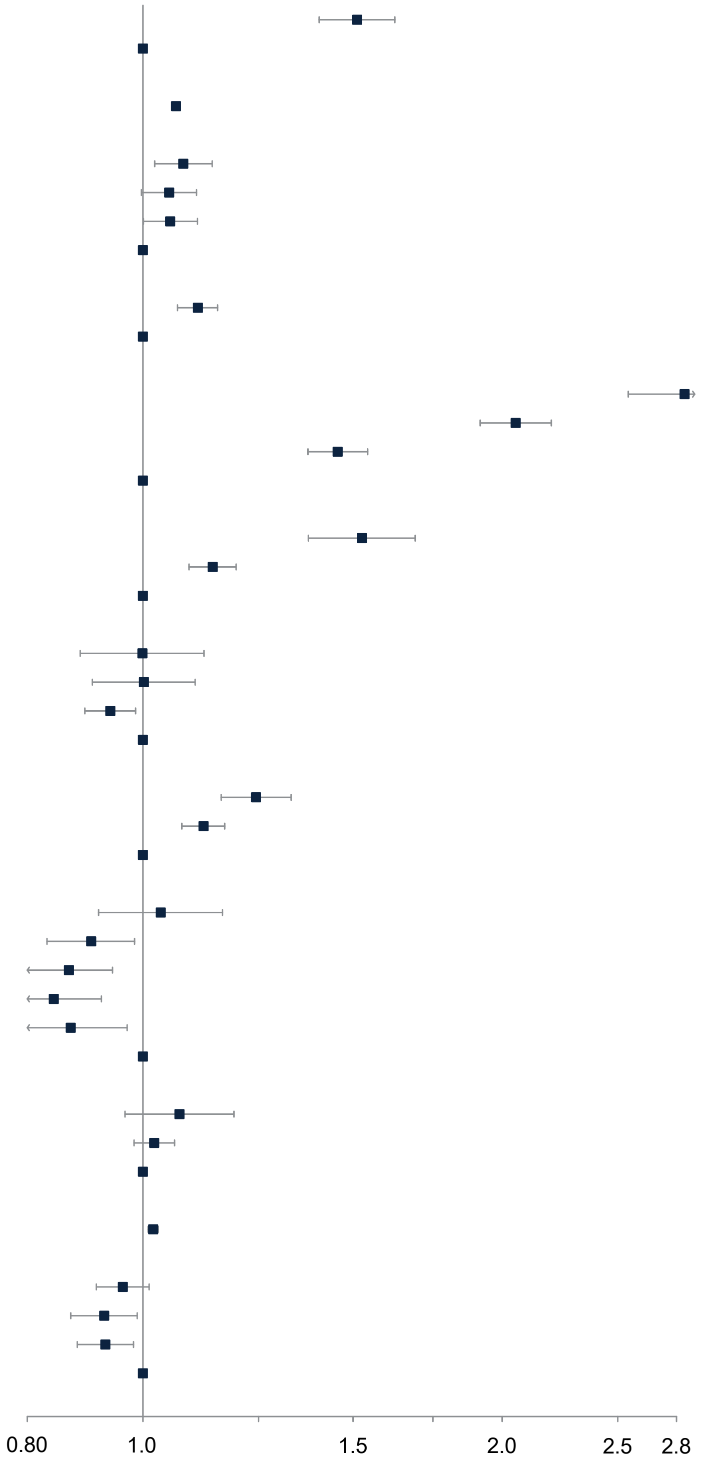

(B) Males with any diagnosed sleep disorders

| Variables | Hazard Ratio (HR) HR [95% CI] |
|---|---|
| **Any Diagnosed Sleep Disorder** | |
| Yes | 1.55 [1.45−1.66] |
| No | 1 |
| **Age of Enrolment** | |
| Each year older | 1.07 [1.06−1.07] |
| **Socioeconomic Status** | |
| Highest Deprivation | 1.04 [0.98−1.09] |
| 2nd Deprivation | 1.03 [0.98−1.08] |
| 3rd Deprivation | 1.01 [0.96−1.06] |
| Least Deprivation | 1 |
| **Mental Health Issues** | |
| Yes | 1.03 [0.99−1.07] |
| No | 1 |
| **Perceived Health** | |
| Poor | 2.16 [1.94−2.4] |
| Fair | 1.7 [1.6−1.81] |
| Good | 1.3 [1.23−1.37] |
| Excellent | 1 |
| **Body Mass Index (kg/m2)** | |
| >= 40 | 1.43 [1.24−1.64] |
| 30 to <40 | 1.16 [1.11−1.2] |
| <30 | 1 |
| **Economic Activity and Shift Work** | |
| Employed in night shift work | 0.99 [0.91−1.07] |
| Employed in day shift work | 1.01 [0.93−1.11] |
| Employed not in shift work | 0.94 [0.9−0.98] |
| Retired/not in the workforce | 1 |
| **Cigarette Smoking** | |
| Current | 1.25 [1.18−1.32] |
| Cessation | 1.09 [1.05−1.13] |
| Never | 1 |
| **Alcohol Drinking** | |
| Quit | 1.15 [0.98−1.34] |
| Occasional | 1.1 [0.96−1.26] |
| < 14 UK Units/wk | 1.02 [0.89−1.16] |
| 14 to <28 UK Units/wk | 0.97 [0.85−1.11] |
| >= 28 UK Units/wk | 1.04 [0.91−1.18] |
| Never | 1 |
| **Dietary Quality** | |
| Poor | 0.99 [0.93−1.06] |
| Intermediate | 1.01 [0.97−1.06] |
| Healthy | 1 |
| **Discretionary Screen Time (hr/d)** | |
| Each hour more | 1.01 [1.01−1.02] |
| **Physical Activity** | |
| No MVPA | 0.92 [0.88−0.97] |
| < 10 MET hrs/wk | 0.95 [0.89−1] |
| 10 to <20 MET hrs/wk | 0.96 [0.91−1.01] |
| >= 20 MET hrs/wk | 1 |

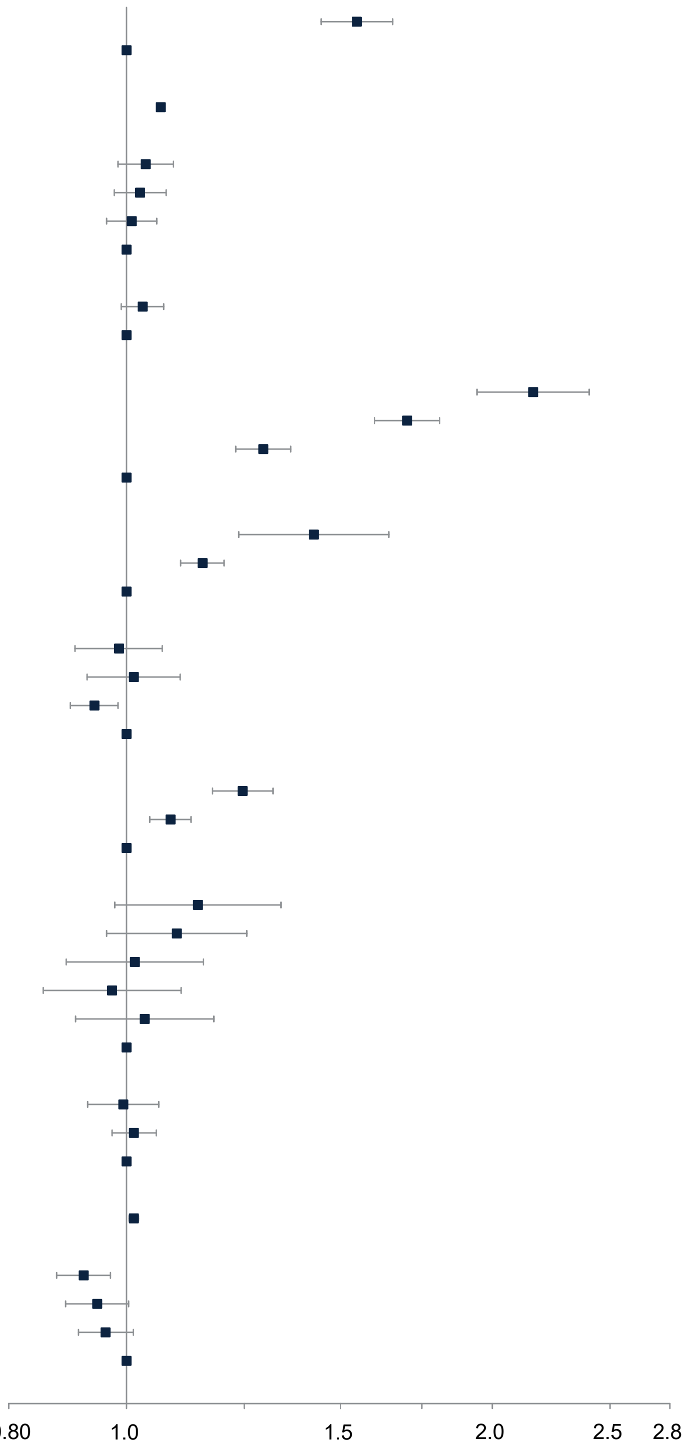

(C) Females with different diagnosed sleep disorders

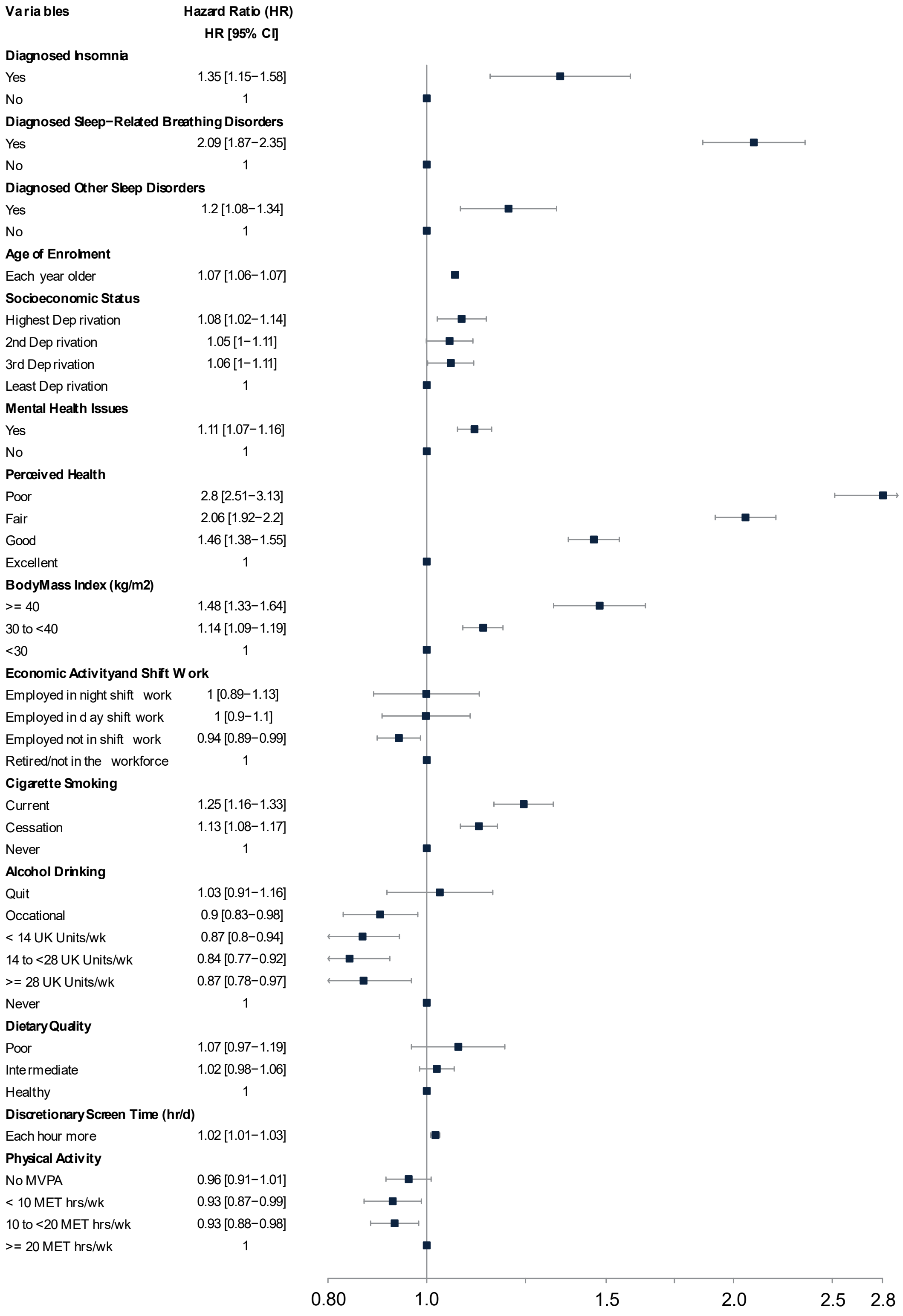

(D) Males with different diagnosed sleep disorders

| Variables | Hazard Ratio (HR) HR [95% CI] |
|---|---|
| **Diagnosed Insomnia** | |
| Yes | 1.49 [1.23−1.8] |
| No | 1 |
| **Diagnosed Sleep−Related Breathing Disorders** | |
| Yes | 1.83 [1.68−1.99] |
| No | 1 |
| **Diagnosed Other Sleep Disorders** | |
| Yes | 1.13 [1−1.28] |
| No | 1 |
| **Age of Enrolment** | |
| Each year older | 1.07 [1.06−1.07] |
| **Socioeconomic Status** | |
| Highest Deprivation | 1.04 [0.98−1.09] |
| 2nd Deprivation | 1.03 [0.98−1.08] |
| 3rd Deprivation | 1.01 [0.96−1.06] |
| Least Deprivation | 1 |
| **Mental Health Issues** | |
| Yes | 1.03 [0.99−1.08] |
| No | 1 |
| **Perceived Health** | |
| Poor | 2.15 [1.94−2.39] |
| Fair | 1.7 [1.6−1.81] |
| Good | 1.3 [1.23−1.37] |
| Excellent | 1 |
| **Body Mass Index (kg/m2)** | |
| >= 40 | 1.37 [1.19−1.58] |
| 30 to <40 | 1.15 [1.1−1.19] |
| <30 | 1 |
| **Economic Activity and Shift Work** | |
| Employed in night shift work | 0.99 [0.91−1.07] |
| Employed in day shift work | 1.02 [0.93−1.11] |
| Employed not in shift work | 0.94 [0.9−0.98] |
| Retired/not in the workforce | 1 |
| **Cigarette Smoking** | |
| Current | 1.25 [1.18−1.32] |
| Cessation | 1.08 [1.04−1.13] |
| Never | 1 |
| **Alcohol Drinking** | |
| Quit | 1.15 [0.98−1.35] |
| Occational | 1.1 [0.96−1.26] |
| < 14 UK Units/wk | 1.02 [0.89−1.16] |
| 14 to <28 UK Units/wk | 0.97 [0.86−1.11] |
| >= 28 UK Units/wk | 1.04 [0.91−1.18] |
| Never | 1 |
| **Dietary Quality** | |
| Poor | 1 [0.93−1.06] |
| Intermediate | 1.01 [0.97−1.06] |
| Healthy | 1 |
| **Discretionary Screen Time (hr/d)** | |
| Each hour more | 1.01 [1.01−1.02] |
| **Physical Activity** | |
| No MVPA | 0.92 [0.87−0.97] |
| < 10 MET hrs/wk | 0.94 [0.89−1] |
| 10 to <20 MET hrs/wk | 0.96 [0.91−1.01] |
| >= 20 MET hrs/wk | 1 |

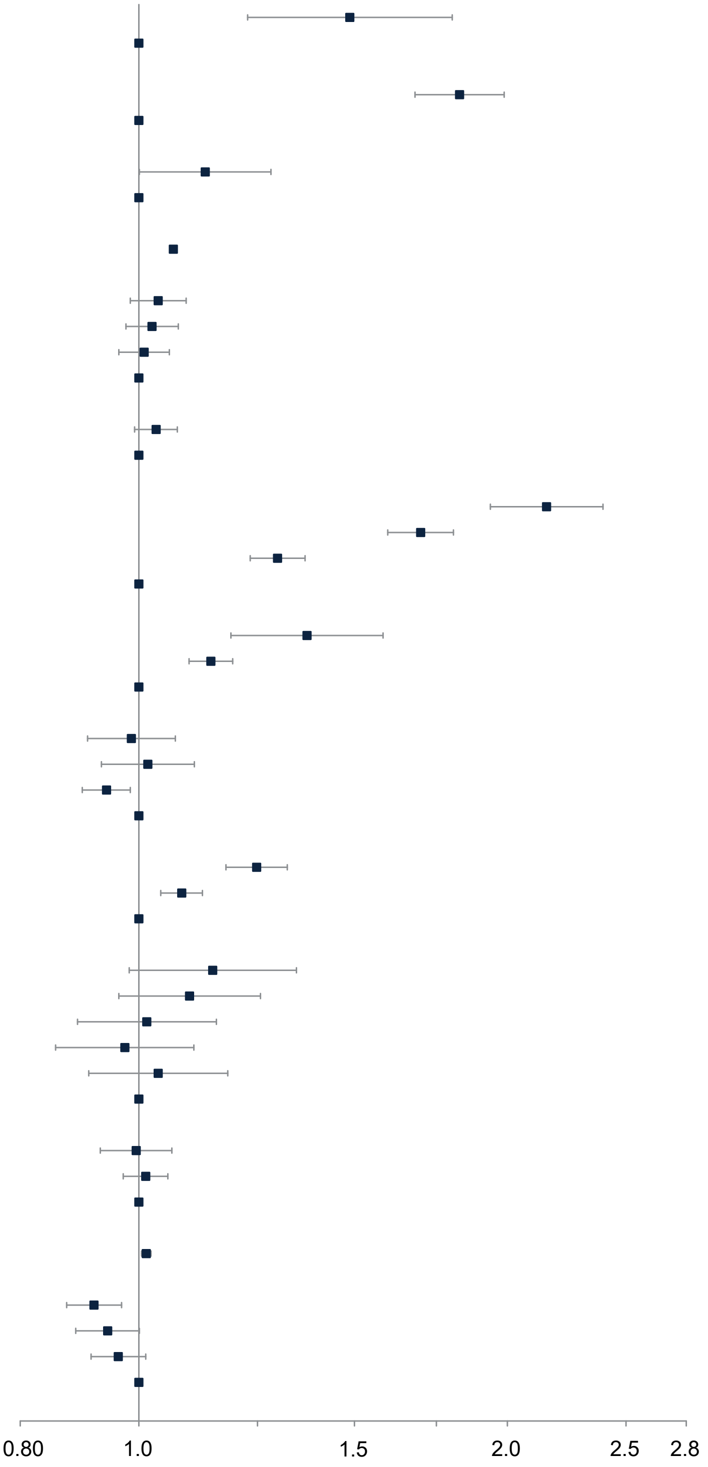
